# Mitochondrial dysfunction underlies cortical atrophy in prodromal synucleinopathies

**DOI:** 10.1101/2022.09.01.22279508

**Authors:** Shady Rahayel, Christina Tremblay, Andrew Vo, Bratislav Mišić, Stéphane Lehéricy, Isabelle Arnulf, Marie Vidailhet, Jean-Christophe Corvol, the ICEBERG Study Group, Jean-François Gagnon, Ronald B. Postuma, Jacques Montplaisir, Simon Lewis, Elie Matar, Kaylena Ehgoetz Martens, Per Borghammer, Karoline Knudsen, Allan K. Hansen, Oury Monchi, Ziv Gan-Or, Alain Dagher

## Abstract

Isolated rapid eye movement sleep behavior disorder (iRBD) is a prodromal synucleinopathy characterized by several changes including brain atrophy. The mechanisms underlying atrophy in iRBD are poorly understood. Here, we performed imaging transcriptomics and comprehensive spatial mapping in a multicentric cohort of 171 polysomnography-confirmed iRBD patients and 238 controls with T1-weighted MRI to investigate the gene expression patterns and the specific neurotransmitter, functional, cytoarchitectonic, and cognitive brain systems associated with cortical thinning in iRBD. We found that genes involved in mitochondrial function and macroautophagy were the strongest contributors to the thinning occurring in iRBD. Moreover, we demonstrated that thinning was constrained by the brain’s connectome and that it mapped onto specific networks involved in motor and planning functions. In contrast with thickness, changes in cortical surface area related to distinct gene and spatial mapping patterns. This study demonstrates that the development of atrophy in synucleinopathies is constrained by specific genes and networks.

## INTRODUCTION

Isolated rapid eye movement sleep behavior disorder (iRBD) is a parasomnia characterized by abnormal movements and vocalizations during REM sleep that typically develops into dementia with Lewy bodies (DLB), Parkinson’s disease (PD), and multiple system atrophy (MSA).^1,2^ As a prodromal synucleinopathy, iRBD patients show brain changes like those seen in manifest synucleinopathies.^3^ In particular, brain atrophy has been shown to occur in iRBD patients,^4^ which correlates with clinical changes,^5-8^ especially cognitive impairment,^6^ and can even predict the phenoconversion to DLB compared to PD.^9^ However, to date, the mechanisms underlying the development of brain atrophy and its patterns in iRBD remain poorly understood.

A collection of observations in humans and experimental findings in animals support the notion that the distribution of pathology in synucleinopathies arises from a prion-like spreading occurring within an environment characterized by some cells being more selectively vulnerable to pathology than others.^10-12^ A recent study tested the prion-like spreading hypothesis and the selective vulnerability hypothesis using an agent-based computational model that simulates (in silico) the propagation of alpha-synuclein based on local gene expression and connectivity.^13^ The simulations derived from this model were shown to recreate the distribution of alpha-synuclein pathology in the mouse brain after the injection of preformed fibrils.^14^ When applied to iRBD and PD, this model demonstrated that the patterns of atrophy observed in these patients could be recreated computationally and that both gene expression and connectivity were determinant factors shaping atrophy.^13,15^ However, in these studies, only the regional expression of *SNCA* and *GBA* was used to model the agents’ spread, thus limiting the breadth of understanding of the wider gene correlates that may underlie structural changes in synucleinopathies. To our knowledge, no study has yet investigated the patterns of gene expression associated with cortical changes in iRBD.

Imaging transcriptomics is a recent approach that allows a multivariate investigation into the associations between brain changes and the transcriptional activity of genes across the whole brain.^16^ In PD, this approach has revealed that the regions showing greater progression of atrophy over two and four years as measured using MRI had a greater expression of genes involved in synaptic activity and cell signaling.^17^ Furthermore, studies computing atrophy spread in PD have also shown the involvement of transcripts associated with immune and lysosomal functions.^18,19^ Imaging transcriptomics has also demonstrated that increased brain iron content in PD as measured using quantitative susceptibility mapping, related to genes implicated in metal detoxification and synaptic transmission/signalling.^20^ In addition, by virtue of arising from a pathological process that spreads through the connectome, many studies have shown that atrophy in neurodegenerative diseases is shaped by connectivity.^13,15,21^ In PD, the progression of atrophy was shown to map onto specific major functional networks, being significantly more pronounced in the limbic, default mode, and visual networks after 2 years, and then extending to almost all networks in the following years.^17^ To our knowledge, the gene expression and connectomics underpinnings of cortical changes in prodromal synucleinopathies remain to be investigated.

Since iRBD is an early stage during which clinical features of synucleinopathies are still modest, identifying abnormalities on MRI scans and understanding their underlying mechanisms may offer new insights into potential therapies aimed at slowing or stopping the neurodegenerative process in patients. In this study, we combined imaging transcriptomics and comprehensive spatial mapping to investigate the genetic and connectivity underpinnings of cortical changes in iRBD. Using a large cohort of polysomnography-confirmed iRBD patients and matched healthy controls with T1-weighted imaging, we applied a partial least squares (PLS) regression approach to identify the gene expression patterns predicting cortical thinning in iRBD and investigated the patterns of biological processes, cellular components, disease-related terms, and cell types enriched in these patterns. We next investigated if the patterns of cortical thinning in iRBD mapped onto specific brain systems, namely neurotransmitter systems, functional networks, cytoarchitectonic classes, and meta-analytic maps of cognitive functions. Next, we assessed whether these patterns were specific to cortical thinning by assessing the gene expression and spatial mapping patterns underlying the changes in cortical surface area in iRBD. We hypothesized that specific gene expression and spatial mapping patterns would underlie cortical thinning in iRBD and that these patterns would be specific to cortical thickness compared to surface area.

## RESULTS

### Demographics and cortical changes in iRBD

Of the 443 participants part of the multicentric cohort,^15^ 34 (7.7%) did not pass deformation-based quality control and 64 (15.6%) did not pass surface-based quality control, resulting in a final sample of 138 patients and 207 controls from which measures of cortical morphometry were derived. All measurements were converted to *W*-scores, which can be thought of as *z*-scores representing deviation from the expected mean of control subjects, while correcting for age, sex, and site.^22,23^ The groups did not differ in age (iRBD: 66.2 ± 7.6, controls: 67.0 ± 6.3, *p*=0.28) and sex (iRBD: 81% men, controls: 77% men, *p*=0.34, see Supp. Table 1 for characteristics of the cohorts). Vertex-wise cortical surface analyses between iRBD patients and controls have been reported elsewhere.^15^ Accounting for age, sex, and site, these showed that iRBD patients have decreased cortical thickness in the left posterior temporal and inferior parietal cortices, the left orbitofrontal and dorsolateral prefrontal cortices, and in the right temporal and lateral occipital cortices (Fig. 1A).^15^ In addition, iRBD patients had increased cortical surface area in the left inferior temporal and entorhinal cortices (Supp. Fig. 1A).^15^ There were no regions with significantly increased cortical thickness or decreased cortical surface area in iRBD patients compared to controls.

**Table 1.** Biological processes enriched in the genes predicting cortical thinning in iRBD.

| GO identifier | GO term | Gene set size | Number of leading edge IDs | Enrichment score | Normalized enrichment score | FDR P-value |
| --- | --- | --- | --- | --- | --- | --- |
| *Negatively weighted (more expressed) genes* |  |  |  |  |  |  |
| GO:0010257 | NADH dehydrogenase complex assembly | 58 | 44 | -0.622 | -2.442 | **<0.0001** |
| GO:0033108 | mitochondrial respiratory chain complex assembly | 88 | 57 | -0.551 | -2.365 | **<0.0001** |
| GO:0072512 | trivalent inorganic cation transport | 32 | 14 | -0.668 | -2.315 | **<0.0001** |
| GO:0009123 | nucleoside monophosphate metabolic process | 275 | 126 | -0.467 | -2.313 | **<0.0001** |
| GO:0070585 | protein localization to mitochondrion | 132 | 53 | -0.492 | -2.227 | **0.00019** |
| GO:0009141 | nucleoside triphosphate metabolic process | 262 | 120 | -0.450 | -2.220 | **0.00016** |
| GO:1902600 | proton transmembrane transport | 121 | 53 | -0.498 | -2.218 | **0.00014** |
| GO:0140053 | mitochondrial gene expression | 158 | 69 | -0.476 | -2.203 | **0.00012** |
| GO:0006091 | generation of precursor metabolites and energy | 405 | 173 | -0.420 | -2.175 | **0.00011** |
| GO:1903008 | organelle disassembly | 93 | 50 | -0.509 | -2.142 | **0.00028** |
| *Positively weighted (less expressed) genes* |  |  |  |  |  |  |
| GO:0022616 | DNA strand elongation | 21 | 11 | 0.638 | 2.081 | **0.015** |
| GO:0002507 | tolerance induction | 13 | 7 | 0.664 | 1.889 | 0.093 |
| GO:0006333 | chromatin assembly or disassembly | 119 | 37 | 0.392 | 1.838 | 0.102 |
| GO:0035329 | hippo signalling | 32 | 19 | 0.486 | 1.777 | 0.140 |
| GO:0008214 | protein dealkylation | 31 | 14 | 0.473 | 1.727 | 0.182 |
| GO:0150076 | neuroinflammatory response | 32 | 12 | 0.472 | 1.709 | 0.176 |
| GO:0033687 | osteoblast proliferation | 21 | 12 | 0.507 | 1.656 | 0.240 |
| GO:0061512 | protein localization to cilium | 39 | 12 | 0.407 | 1.550 | 0.265 |
| GO:0010463 | mesenchymal cell proliferation | 34 | 13 | 0.430 | 1.548 | 0.252 |
| GO:0042490 | mechanoreceptor differentiation | 44 | 15 | 0.380 | 1.517 | 0.267 |

The top 10 biological process terms enriched in the genes predicting cortical thinning in iRBD are reported ranked based on the normalized enrichment score. Bold values represent the terms significantly enriched after applying FDR correction.
FDR = false discovery rate; GO = Gene Ontology; GSEA = gene set enrichment analysis; iRBD = isolated REM sleep behavior disorder; PML = promyelocytic leukemia; REM = rapid eye movement.

**Figure 1.**
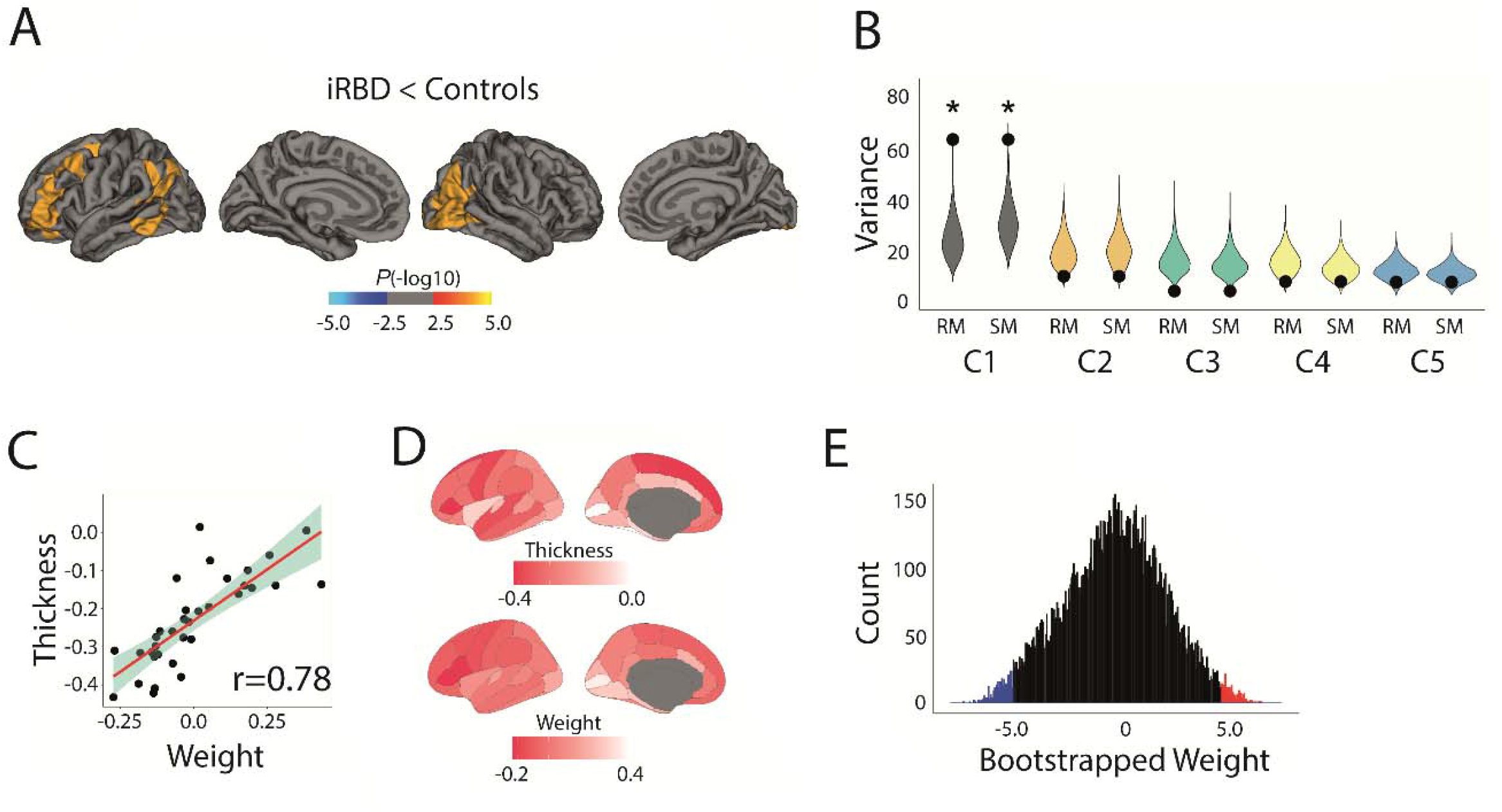
Patterns of gene expression underlying cortical thinning in iRBD. **(A)** Vertex-wise patterns showing the significant changes in cortical thickness in iRBD patients compared to controls. **(B)** Violin plots showing the percentage of variance in cortical thickness *W*-scores explained by gene expression; the dot represents the empirical variance, and the asterisk indicates the components that were significant against random and spatial null models. **(C)** Scatterplots of the associations between thickness *W*-scores and the regional weights of the first components. **(D)** Brain renderings of the thickness *W*-scores and the regional weights of the first components. **(E)** Density plots of each gene’s bootstrapped weight on the first components; gene set enrichment analysis was performed on all genes, whereas over-representation analysis was performed on genes with bootstrap ratios ± 5.0. C = component; iRBD = isolated REM sleep behavior disorder; REM = rapid eye movement; RM = random null models; SM = spatial null models.

### Cortical thinning associates with mitochondrial function

PLS regression was performed to compare spatial patterns of gene expression versus cortical thinning in iRBD. Of the five components tested, only the first explained significantly more variance in thickness than random null models (60.7% versus 23.4%, *p*_random_<0.0001) and spatial null models that preserved the spatial autocorrelation between brain regions (60.7% versus 29.9%, *p*_spatial_=0.0005) (Fig. 1B). The regional weights of the significant component were positively associated with cortical thickness *W*-scores in iRBD patients (*r*=0.78, *p*<0.0001) (Fig. 1C), meaning that the genes negatively weighted on the component were more expressed in regions with greater cortical thinning and that the genes positively weighted on the component were less expressed in regions with greater cortical thinning (Fig. 1D).

We next applied gene set enrichment analysis (GSEA) to examine the translational relevance of the genes whose expression overlapped with cortical thickness changes in iRBD. The genes were ranked based on bootstrap ratio (Fig. 1E), and the resulting ranked gene list was intersected with several knowledge bases to identify if specific biological processes, cellular components, and disease-related terms were enriched at the top (positively weighted genes) or at the bottom (negatively weighted genes) of the list. In terms of biological processes, GSEA revealed that the genes most strongly expressed in association with greater cortical thinning (i.e., the negatively weighted genes on the component) were enriched for processes involved in oxidative phosphorylation, with NADH dehydrogenase complex (complex I) assembly (normalized enrichment score: -2.44, *P*_*FDR*_<0.0001), mitochondrial respiratory chain complex assembly (−2.37, *P*_*FDR*_<0.0001), and trivalent inorganic cation transport (−2.32, *P*_*FDR*_<0.0001) ranking as the most strongly enriched terms (Fig. 2A and Table 1). Of the 15,633 genes used as input to PLS regression, 554 (3.5%) genes were robustly associated with thickness *W*-scores in iRBD (i.e., bootstrap ratio weight ±5.0), with 332 (60%) being negatively weighted and 222 (40%) being positively weighted (Fig. 1E). When assessing the enrichment of these genes compared to the complete gene list, over-representation analysis showed that the genes most strongly expressed in association with cortical thinning were additionally enriched for processes involved in macroautophagy (Fig. 2B).

**Figure 2.**
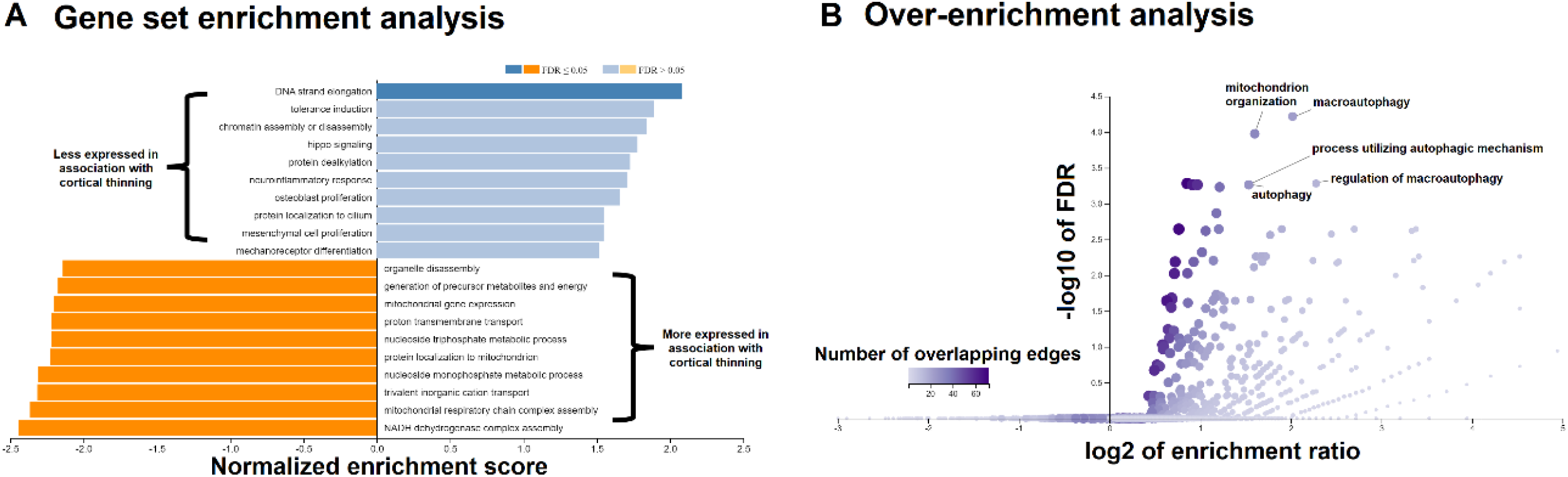
Enrichment analyses of the genes associated with cortical thinning in iRBD. **(A)** The top 10 biological process terms from the GO Consortium knowledge base that are enriched in the positively and negatively weighted gene sets predicting cortical thinning in iRBD. Terms are ranked based on the normalized enrichment score; darker colored bars present significantly enriched terms after FDR correction. **(B)** Volcano plots of the over-representation analyses showing the biological process terms enriched in the genes most strongly associated with cortical thinning in iRBD (bootstrap ratio <-5.0). The color bar represents the number of overlapping edges for each gene category and the size of the dot represents the size of the gene category. FDR = false discovery rate; GO = Gene Ontology; iRBD = isolated REM sleep behavior disorder; REM = rapid eye movement.

In terms of cellular components, GSEA of the ranked gene list showed that the genes more expressed in association with cortical thinning were significantly enriched for components localized to the mitochondrion, including the mitochondrial membrane (−2.84, *P*_*FDR*_<0.0001), the respiratory chain (−2.80, *P*_*FDR*_<0.0001), the mitochondrial protein complex (−2.72, *P*_*FDR*_<0.0001), and the NADH dehydrogenase complex (or complex I) (−2.65, *P*_*FDR*_<0.0001) (Table 2). In terms of disease-related terms, when using the DisGeNET, OMIM, and PharmGKB disease databases, the genes more expressed with cortical thinning in iRBD were enriched for terms related to mitochondrial diseases and lactic acidosis (Table 3).

**Table 2.** Cellular components enriched in the genes predicting cortical thinning in iRBD.

| GO identifier | GO term | Gene set size | Number of leading edge IDs | Enrichment score | Normalized enrichment score | FDR P-value |
| --- | --- | --- | --- | --- | --- | --- |
| *Negatively weighted (more expressed) genes* |  |  |  |  |  |  |
| *Cellular components* |  |  |  |  |  |  |
| GO:0044455 | mitochondrial membrane part | 206 | 128 | -0.589 | -2.843 | **<0.0001** |
| GO:0070469 | respiratory chain | 86 | 61 | -0.656 | -2.799 | **<0.0001** |
| GO:0098798 | mitochondrial protein complex | 248 | 135 | -0.554 | -2.724 | **<0.0001** |
| GO:0030964 | NADH dehydrogenase complex | 45 | 35 | -0.699 | -2.645 | **<0.0001** |
| GO:0005743 | mitochondrial inner membrane | 414 | 200 | -0.501 | -2.596 | **<0.0001** |
| GO:0016469 | proton-transporting two-sector ATPase complex | 40 | 22 | -0.689 | -2.497 | **<0.0001** |
| GO:0070069 | cytochrome complex | 27 | 17 | -0.682 | -2.258 | **<0.0001** |
| GO:1905368 | peptidase complex | 85 | 37 | -0.526 | -2.230 | **<0.0001** |
| GO:0005759 | mitochondrial matrix | 432 | 183 | -0.428 | -2.230 | **<0.0001** |
| GO:0043209 | myelin sheath | 152 | 75 | -0.473 | -2.179 | **<0.0001** |
| *Positively weighted (less expressed) genes* |  |  |  |  |  |  |
| *Cellular components* |  |  |  |  |  |  |
| GO:0044815 | DNA packaging complex | 52 | 34 | 0.598 | 2.416 | **<0.0001** |
| GO:0032993 | protein-DNA complex | 129 | 46 | 0.400 | 1.911 | **0.012** |
| GO:0032039 | integrator complex | 17 | 6 | 0.611 | 1.900 | **0.009** |
| GO:0000790 | nuclear chromatin | 297 | 111 | 0.320 | 1.704 | **0.043** |
| GO:0017053 | transcriptional repressor complex | 78 | 27 | 0.384 | 1.693 | **0.037** |
| GO:0005790 | smooth endoplasmic reticulum | 31 | 16 | 0.445 | 1.602 | 0.068 |
| GO:0044450 | microtubule organizing center part | 155 | 38 | 0.325 | 1.588 | 0.064 |
| GO:0016605 | PML body | 91 | 20 | 0.335 | 1.514 | 0.079 |
| GO:0005697 | telomerase holoenzyme complex | 19 | 9 | 0.477 | 1.502 | 0.079 |
| GO:0045178 | basal part of cell | 41 | 19 | 0.395 | 1.501 | 0.073 |

The top 10 biological process and cellular component terms enriched in the genes predicting cortical thickness changes in iRBD are reported ranked based on the normalized enrichment score. Bold values represent the terms significantly enriched after applying FDR correction.
FDR = false discovery rate; GO = Gene Ontology; GSEA = gene set enrichment analysis; iRBD = isolated REM sleep behavior disorder; PML = promyelocytic leukemia; REM = rapid eye movement.

**Table 3.** Disease-related terms enriched in the genes predicting cortical thinning in iRBD.

| Term | Description | Gene set size | Number of leading edge IDs | Enrichment score | Normalized enrichment score | FDR *P*-value |
| --- | --- | --- | --- | --- | --- | --- |
| *Negatively weighted (more expressed) genes* |  |  |  |  |  |  |
| *DisGeNET* |  |  |  |  |  |  |
| C0001125 | Acidosis, Lactic | 87 | 48 | -0.60643 | -2.5857 | **<0.0001** |
| C0347959 | Lactic acidemia | 85 | 47 | -0.60161 | -2.5503 | **<0.0001** |
| C1167918 | CSF lactate increased | 33 | 22 | -0.68981 | -2.4097 | **<0.0001** |
| C0006114 | Cerebral edema | 23 | 15 | -0.70202 | -2.2295 | **<0.001** |
| C0023264 | Leigh disease | 33 | 21 | -0.62946 | -2.2161 | **<0.001** |
| C0424551 | Impaired exercise tolerance | 42 | 22 | -0.59938 | -2.2105 | **<0.001** |
| C1836440 | Increased serum lactate | 52 | 27 | -0.57067 | -2.1782 | **0.0012** |
| C4021546 | Abnormal mitochondria in muscle tissue | 17 | 12 | -0.75098 | -2.1770 | **0.0011** |
| C1145670 | Respiratory failure | 73 | 40 | -0.52822 | -2.1488 | **0.0017** |
| C1855020 | Acute necrotizing encephalopathy | 14 | 11 | -0.77116 | -2.1256 | **0.0027** |
| *OMIM* |  |  |  |  |  |  |
| 252010 | Mitochondrial complex I deficiency | 20 | 15 | -0.63990 | -2.0187 | **<0.001** |
| *PharmGKB* |  |  |  |  |  |  |
| PA447172 | Mitochondrial diseases | 349 | 161 | -0.46404 | -2.3516 | **<0.0001** |
| PA166048819 | Acidosis, Respiratory | 78 | 49 | -0.55240 | -2.2995 | **<0.0001** |
| PA443242 | Acidosis | 117 | 66 | -0.51360 | -2.2808 | **<0.0001** |
| PA445837 | Thiamine deficiency | 54 | 25 | -0.57049 | -2.2013 | **0.0012** |
| PA165108683 | Pyruvate dehydrogenase complex deficiency | 26 | 12 | -0.65683 | -2.1974 | **<0.001** |
| PA443243 | Acidosis, Lactic | 64 | 24 | -0.54429 | -2.1955 | **<0.001** |
| PA447190 | Cytochrome-c oxidase deficiency | 55 | 30 | -0.55467 | -2.1675 | **0.0016** |
| PA446467 | Mitochondrial encephalomyopathies | 48 | 24 | -0.57178 | -2.1459 | **0.0018** |
| PA165857066 | Anemia, Hemolytic, Congenital Nonspherocytic | 10 | 7 | -0.82012 | -2.0815 | **0.0062** |
| PA165108373 | Biotinidase deficiency | 44 | 22 | -0.55813 | -2.0769 | **0.0058** |

The human disease terms enriched in the genes predicting cortical thinning in iRBD using the DisGeNET, OMIM, and PharmGKB knowledge bases. Terms are reported ranked based on the normalized enrichment score. Only terms enriched after applying FDR correction are shown.
FDR = false discovery rate; GO = Gene Ontology; GSEA = gene set enrichment analysis; iRBD = isolated REM sleep behavior disorder; PML = promyelocytic leukemia; REM = rapid eye movement.

In contrast, the genes less strongly expressed in association with greater cortical thinning (i.e., the positively weighted genes on the component) were enriched for DNA strand elongation (normalized enrichment score: 2.08, *P*_*FDR*_=0.015) (Fig. 2A and Table 1) and localized to nuclear-related cellular components, namely the DNA packaging complex (2.42, *P*_*FDR*_<0.0001), the protein-DNA complex (1.91, *P*_*FDR*_=0.012), the integrator complex (1.90, *P*_*FDR*_=0.009), the nuclear chromatin (1.70, *P*_*FDR*_=0.043), and the transcriptional repressor complex (1.69, *P*_*FDR*_=0.037) (Fig. 2A and Table 2). No disease-related gene terms were significantly enriched for these genes.

Using single-cell mRNA sequencing data from post-mortem human brain samples,^24^ we performed a virtual histology approach to investigate whether cortical thinning in iRBD was associated with the expression of genes corresponding to specific cell types across the cortex, namely astrocytes, endothelial cells, microglia, excitatory and inhibitory neurons, oligodendrocytes, and oligodendrocyte precursor cells. We found that the pattern of cortical thickness *W*-scores in iRBD patients did not associate significantly with the gene expression of any of the 7 cell types (Fig. 3 and Supp. Table 2).

**Figure 3.**
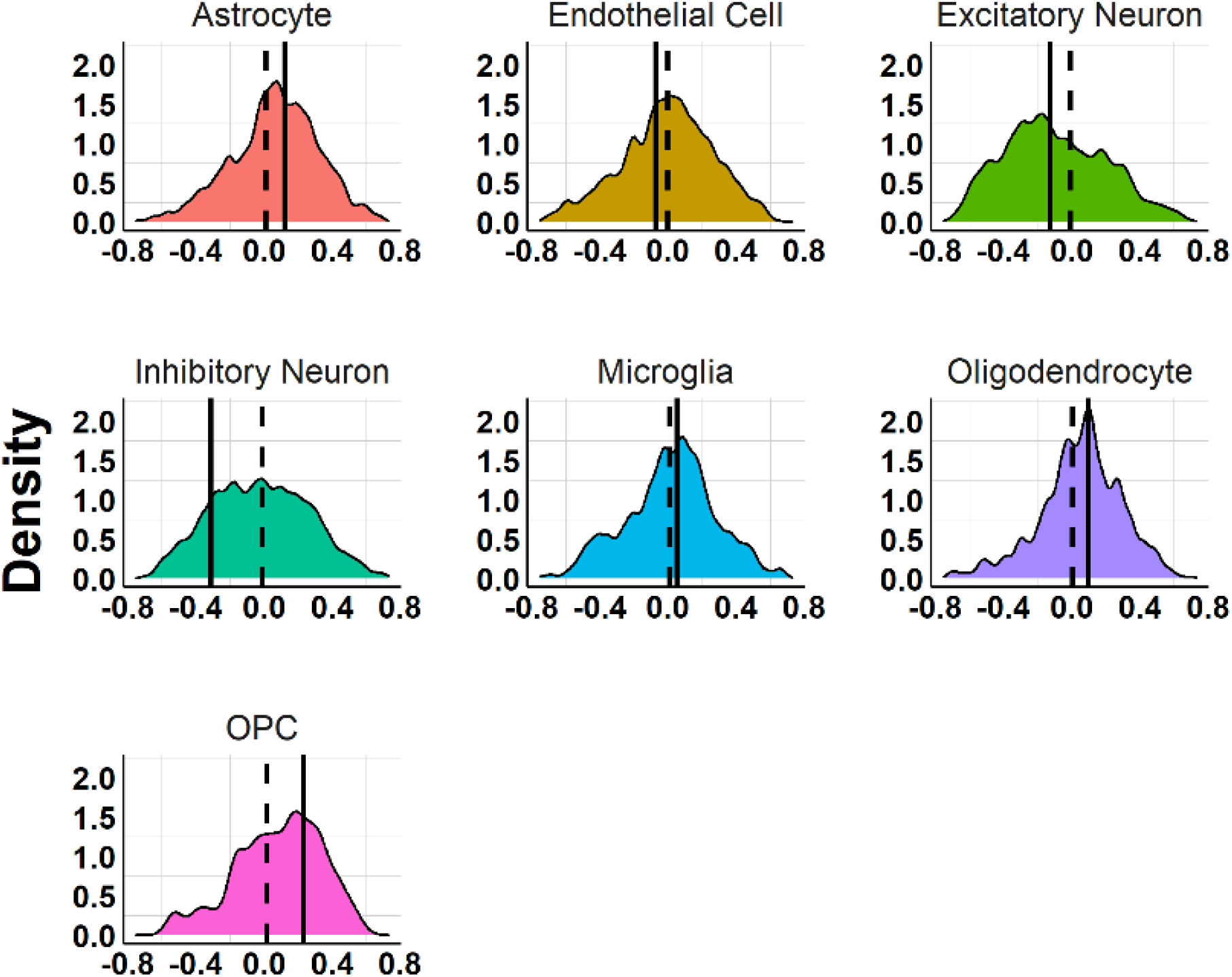
Relationship between cell type gene expression and cortical thinning in iRBD. Density plots showing the distribution of the correlation coefficients between regional cortical thickness *W*-scores and the average regional expression of genes associated with each of the 7 cell types. The straight line represents the empirical correlation, and the dashed line represents the average correlation coefficient observed in sets of 10,000 spatially constrained null models. None of the associations was significant after correcting for multiple comparisons. iRBD = isolated REM sleep behavior disorder; OPC = oligodendrocyte precursor cells; REM = rapid eye movement.

To ensure that our patterns were not due to the gene ontology database chosen, we repeated the analyses using the GOrilla platform and assessed the biological processes and cellular components enriched in the same gene lists. In line with our findings using the WebGestalt 2019 platform, genes more expressed in association with cortical thinning in iRBD were enriched for the regulation of macroautophagy and mitochondrial structures, whereas genes less expressed with cortical thinning were enriched for RNA processing and the nucleoplasm (Supp. Tables 3 and 4).

### The connectome constrains cortical changes in iRBD

A diverse collection of clinical, experimental, and computational studies support that alpha-synuclein pathology behaves in a prion-like fashion,^10,12,13,25-27^ including in iRBD.^15^ Here, we used a structural and functional neighborhood analysis to investigate whether the patterns of cortical thinning in iRBD were constrained by the structural and functional connectivity architecture of the brain. We tested whether the thickness *W*-score of each region was dependent on the average thickness *W*-scores of its connected neighbors. We found that the greater the cortical thinning in a region in iRBD, the greater the thinning in the neighborhood of regions sharing a structural (*r*=0.55, *p*_spatial_=0.0132, *p*_random_<0.0001) or functional connection (*r*=0.52, *p*_spatial_=0.0108, *p*_random_<0.0001) (Fig. 4).

**Figure 4.**
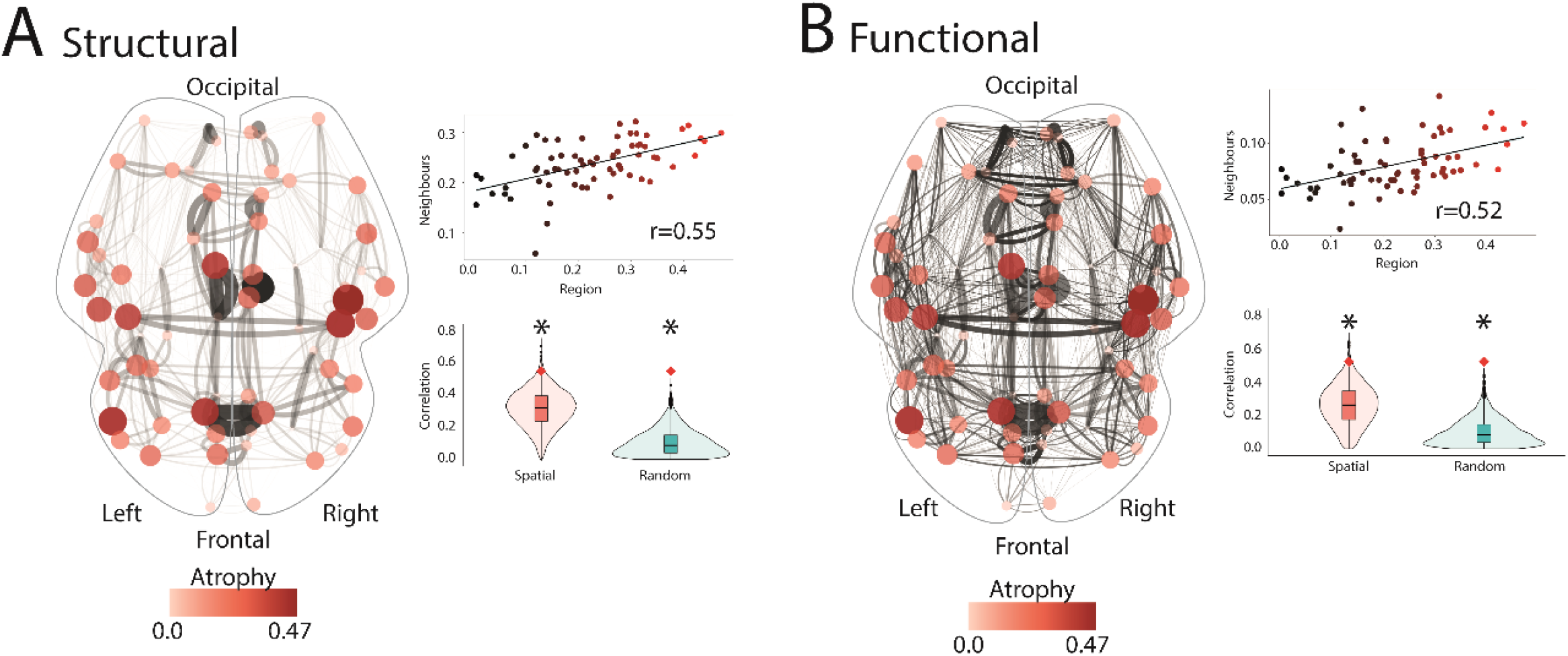
The connectome constrains cortical thinning in iRBD. Brain renderings showing the associations between the deviation in cortical thickness *W*-scores in iRBD and **(A)** structural and **(B)** functional connectivity. The edge thickness represents the interregional connection strength, whereas the node size and color represent the local deviation in the *W*-score in iRBD compared to controls (i.e., the larger and redder, the larger the thinning). The scatterplots show the associations between the deviations in *W*-scores and the average *W*-scores observed in structural or functional neighbors. The violin plots show the empirical correlation against sets of 10,000 correlations generated from spatial and random null models. The asterisk indicates associations that were significant against null models. iRBD = isolated REM sleep behavior disorder; REM = rapid eye movement.

### Cortical thinning in iRBD is associated with DAT density

We next sought to determine whether the pattern of cortical changes in iRBD associated with specific neurotransmitter systems. First, we tested the relationships between the cortical thickness *W*-scores and the regional tracer density values of 18 receptors, transporters, and receptor binding sites associated with dopamine, serotonin, noradrenaline, acetylcholine, GABA, glutamate, histamine, cannabinoids, and opioids.^28^ We found that regions showing cortical thinning in iRBD had a lower density of DAT (*r*=0.51, *p*_spatial_=0.0008, *p*_random_<0.0001), 5-HTT (*r*=0.51, *p*_spatial_=0.003, *p*_random_=<0.0001) and D_1_ (*r*=0.39, *p*_spatial_=0.014, *p*_random_=0.0003), and a higher density of NET (*r*=-0.36, *p*_spatial_=0.041, *p*_random_=0.002) (Fig. 5); however, only the DAT association was significant after Bonferroni correction.

**Figure 5.**
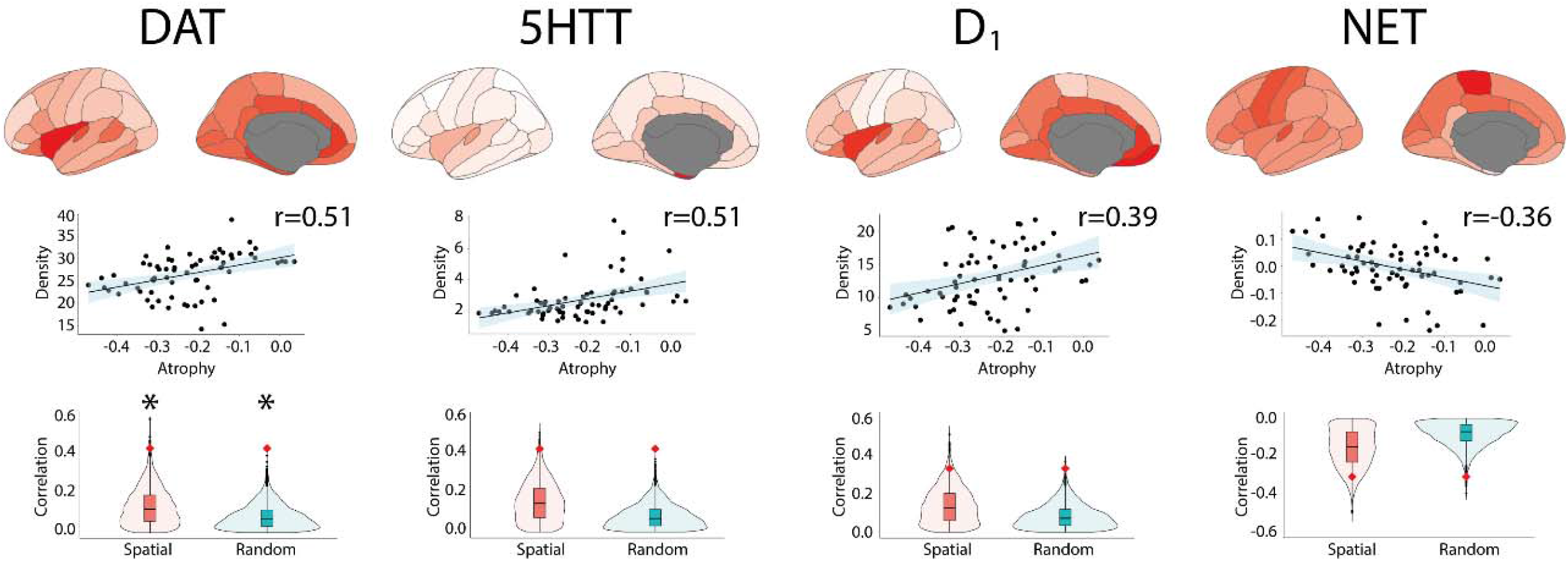
Cortical thinning in iRBD map onto regional tracer density. Brain renderings and scatterplots showing the tracer density maps of the receptors, transporters, and binding sites associated with cortical thickness *W*-scores in iRBD. The violin plots show the empirical correlations tested against distributions of correlations from sets of spatial and random null models. The asterisk indicates the significant associations after Bonferroni correction. iRBD = isolated REM sleep behavior disorder; REM = rapid eye movement.

### Cortical thinning in iRBD maps onto the motor system

We next investigated whether the patterns of cortical thickness *W*-scores in iRBD mapped onto specific resting-state networks, cytoarchitectonic classes, and functional correlates of cognition. The correspondence with resting-state networks was assessed using the 7-network parcellation by Yeo et al.,^29^ which assigns regions to the visual, somatomotor, dorsal attention, ventral attention, limbic, frontoparietal, and default-mode networks. We found that cortical thinning in iRBD was more pronounced in the somatomotor (average atrophy: -0.32, *p*_spatial_=0.017, *p*_random_=0.001) and default-mode networks (−0.27, *p*_spatial_=0.033, *p*_random_=0.036) (Fig. 6A). The correspondence with cytoarchitectonic classes was assessed using the extended version of the von Economo and Koskinas atlas,^30,31^ which comprises the primary motor cortex, association cortices, primary and secondary sensory areas, limbic, and insular areas. We found that cortical thinning in iRBD was significantly more pronounced in the primary motor cortex (−0.39, *p*_spatial_=0.006, *p*_random_=0.0009) (Fig. 6B). The correspondence between the cortical thickness and surface area *W*-scores in iRBD and the functional correlates of 123 cognitive processes also revealed that greater cortical thinning was found in regions more activated during tasks related to planning (*r*=-0.48, *p*_spatial_=0.010, *p*_random_<0.0001) and action (*r*=-0.45, *p*_spatial_=0.026, *p*_random_<0.0001).

**Figure 6.**
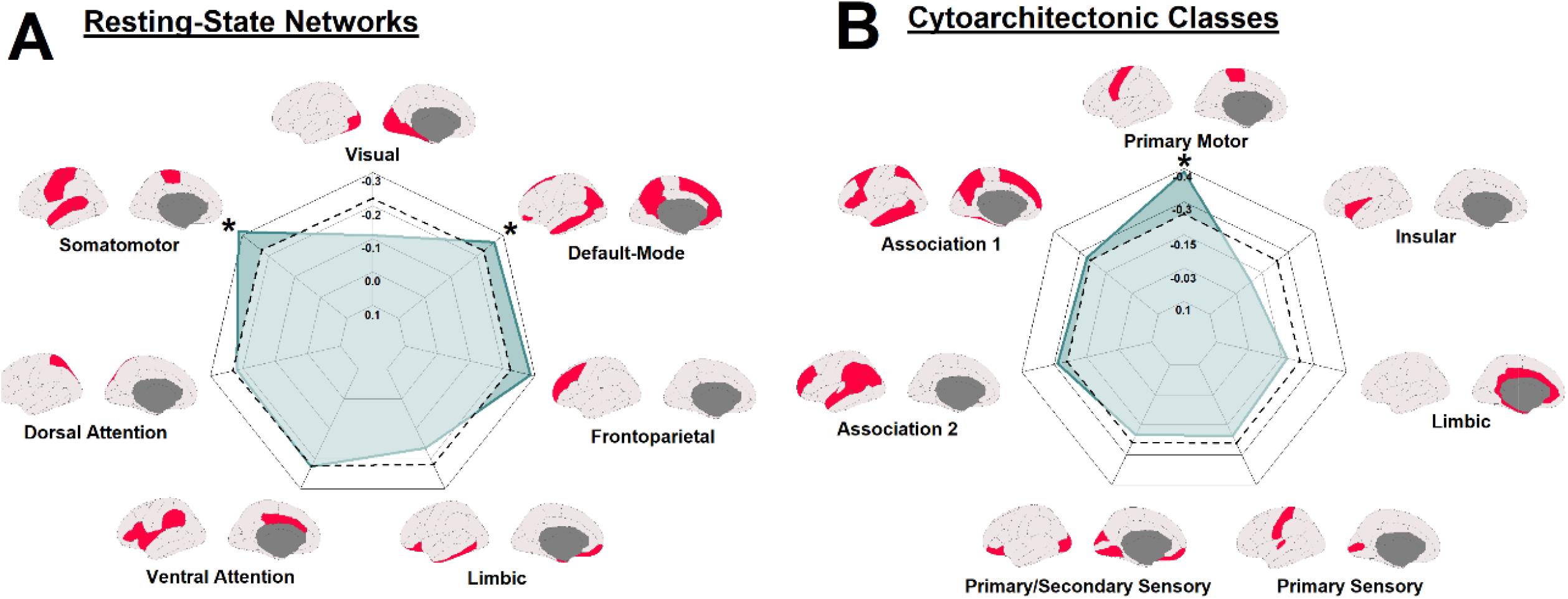
Cortical thinning in iRBD map onto specific resting-state networks and cytoarchitectonic classes. Radar charts showing the correlation between cortical thickness *W*-scores in iRBD and **(A)** resting-state networks and **(B)** cytoarchitectonic classes. The regular line represents the empirical correlations, and the dashed line represents the average correlation observed in sets of 10,000 spatial null models. The asterisk indicates the networks and classes where the observed spatial correlation was significantly different from the null correlation. iRBD = isolated REM sleep behavior disorder; REM = rapid eye movement.

### Cortical thinning and surface area have distinct underlying patterns

Most large-scale studies of human cortical morphometry find that cortical thickness and surface area measured by MRI are dissociable and independent.^32-34^ To investigate whether the gene and spatial patterns were specific to cortical thinning, we applied the same analyses to cortical surface area *W*-scores in iRBD. For the gene expression analyses, PLS regression identified one component that explained significantly more variance in cortical surface area *W*-scores than null models (random null: 42.7% versus 23.4%, *p*_random_=0.015; spatial null: 42.7% versus 26.3%, *p*_spatial_=0.036) (Supp. Fig. 1B). Positively weighted genes on the component were more expressed in association with greater surface area, whereas negatively weighed genes were less expressed in association with greater surface area (Supp. Fig. 1C and 1D). The positively weighted genes were enriched for processes involved in the inflammatory response and metal detoxification, whereas the negatively weighted genes were not enriched for any biological process or cellular component gene term (Supp. Fig 2 and Supp. Table 5). These enrichment patterns were similar for both gene enrichment platform (Supp. Tables 3 and 4). When applying virtual histology, in contrast to cortical thickness, we found that greater regional cortical surface area occurred in regions with a greater expression of genes specific to astrocytes (*r*=0.61, *p*_spatial_=0.0011, *p*_random_=0.0001), microglia (*r*=0.52, *p*_spatial_=0.0053, *p*_random_=0.001), and oligodendrocyte precursor cells (*r*=0.53, *p*_spatial_=0.0049, *p*_random_=0.0007) (Supp. Fig. 3 and Supp. Table 6).

For the connectivity and spatial mapping analyses, the structural and functional neighborhood analysis showed that the regional cortical surface area *W*-scores in iRBD were positively associated with the change in surface area in the regions sharing a structural or functional connection (Supp. Fig 4), supporting that the connectome also constrains the surface area changes in iRBD. We also found that greater cortical surface area in iRBD was related to a lower density of GABA_A/BZ_, 5-HT_6_, NET, 5-HT_1B_ and M_1_, and to a higher density of 5-HT_1A_ and 5-HT_4_ (Supp. Fig. 5); however, only GABA_A/BZ_ remained significant after multiple comparison correction. Finally, we found that cortical surface area was decreased in the visual resting-state network and increased in the limbic resting-state network (Supp. Fig. 6A). Cytoarchitectonically, cortical surface area was decreased in the primary and secondary sensory areas and increased in the limbic areas (Supp. Fig. 6B). Decreased cortical surface area in iRBD associated with regions more activated during tasks related to gaze and fixation (Supp. Fig. 7). Collectively, this demonstrates that cortical thinning in iRBD is associated with gene expression and spatial mapping patterns that are different from those underlying changes in cortical surface area.

**Figure 7.**
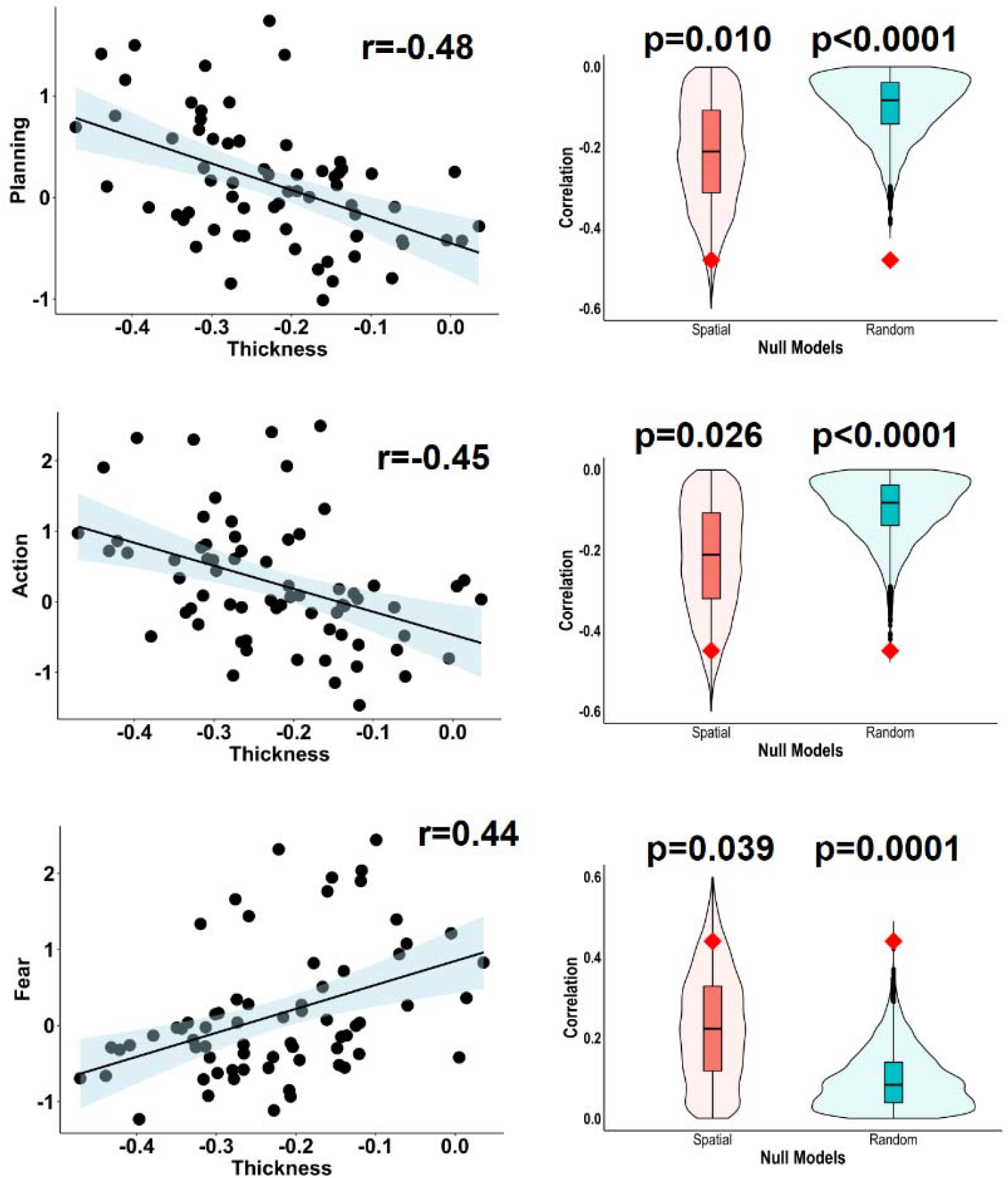
Cortical thinning in iRBD map onto specific correlates of cognitive processes. Scatterplots showing the associations between cortical thickness *W*-scores in iRBD and the functional correlates of 123 different cognitive processes. The violin plots show the empirical correlations compared against the distributions of correlations from sets of 10,000 spatial and random null models. Of the 123 processes tested, only 3 were significant following Bonferroni correction. iRBD = isolated REM sleep behavior disorder; REM = rapid eye movement.

## DISCUSSION

Patients with iRBD demonstrate brain atrophy even prior to the development of motor and cognitive symptoms associated with synucleinopathy.^4-6,8,9,15,35,36^ Brain atrophy in iRBD is associated with motor and cognitive features, predicts conversion towards dementia, and appears to reflect the spread of abnormal alpha-synuclein isoforms in the brain.^5,6,8,9,15,37^ To better understand the pathophysiology of iRBD, we used an imaging transcriptomics approach with comprehensive spatial mapping to investigate the gene correlates and the spatial patterning of cortical thinning in iRBD. We found that cortical thinning is associated with mitochondrial function and macroautophagy and that the pattern of thinning is constrained by the structural and functional architecture of the connectome. We further demonstrated that the pattern of thinning maps onto specific brain systems related to the motor and planning functions. We also showed that these gene and spatial mapping patterns were specific to cortical atrophy in iRBD in that changes in cortical surface area in the same patients related to different genes and brain networks. Altogether, this study provides insight into the processes underlying the development of cortical atrophy in synucleinopathies.

Synucleinopathies are associated with several pathogenic events, with mitochondrial dysfunction, abnormal protein degradation, and inflammation being among the most reported.^38,39^ In this study, we found that cortical thinning in iRBD was increased in regions with a greater expression of genes involved in mitochondrial functioning, particularly in the NADH dehydrogenase complex or complex I. Complex I deficiency in the substantia nigra is a hallmark of PD^40^ and is found in the brain and peripheral tissues of PD patients.^41-43^ In the present study, the genes that were the most robustly associated with cortical thinning in iRBD (i.e., most negative bootstrap ratio weights) were also enriched for macroautophagy, a major autophagic pathway that engulfs material to be degraded into autophagosomes before fusing with lysosomes for degradation.^44^ A previous study in PD that used imaging transcriptomics to assess the gene expression correlates of brain atrophy progression over 4 years found that the regions with the greatest atrophy progression were enriched for genes involved in synaptic transmission and cell signaling.^17^ Taken together, this supports the sequence of events by which pathologic alpha-synuclein first interferes with mitochondrial functioning, then leads to synaptic dysfunctioning,^45-47^ and eventually manifests as cortical thinning in patients, including during the prodromal period.

In this study, we also found that the pattern of cortical thinning in iRBD followed the constraints imposed by the brain’s structural and functional architecture. According to the prion-like spread hypothesis, alpha-synuclein pathology propagates between cells and imposes its abnormal template onto native alpha-synuclein proteins, amplifying the pathological process.^48^ We recently provided evidence in favor of this hypothesis using a computational spreading model of alpha-synuclein (agent-based Susceptible-Infected-Removed (SIR) model), showing that the distribution of alpha-synuclein pathology quantified in mice injected with preformed fibrils could be recreated based on connectivity and gene expression. Importantly, the SIR model was also able to recreate the atrophy patterns observed in iRBD and PD, supporting the prion-like spread model in synucleinopathies.^13,15^ The constraining effect of both the structural and functional connectome on atrophy in iRBD is in line with this. A logical corollary of this spreading hypothesis is that if alpha-synuclein pathology spreads via connections and that connectivity constrains the atrophy in iRBD, then it is expected that the atrophy in iRBD maps onto specific brain systems. This is what we observed: the pattern of cortical thinning in iRBD was significantly more pronounced in the motor and default-mode networks, whereas the pattern of cortical surface area changes in iRBD was more pronounced in limbic and visual networks. These networks are in line with the spatial patterning of atrophy progression reported in PD over 2 years,^49^ anchoring further our atrophy pattern within the prodromal spectrum of synucleinopathies.

Our findings revealed the presence of a dichotomy in the underlying genetic and connectomics bases of cortical thinning and cortical surface area changes in iRBD. Unlike cortical thickness where thinning is generally the event expected from aging and neurodegeneration,^50^ surface area was increased in certain areas compared to healthy individuals.^15^ This is similar to a population-based MRI study of healthy adults showing that a higher genetic risk of PD is associated with greater cortical surface area.^51^ In our study, we showed that greater cortical surface area in iRBD was associated with a greater expression of genes involved in the inflammatory response. Similarly, our virtual histology analysis showed that increased surface area related to genes associated with astrocytes and microglia. These findings are in line with a large body of evidence that supports a link between inflammation and synucleinopathies, particularly in the prodromal stages,^38^ where the activation of toxic microglia is thought to lead to the production of proinflammatory cytokines, which in turn produce cellular damage and cell death. A recent investigation confirmed this sequence of events, showing that pro-inflammatory cytokines are a causal factor of prodromal PD and that an earlier age at onset of PD is associated with increased interleukin-6 concentration.^52^ In addition, we found that greater surface area in iRBD occurred in regions with a greater expression of genes involved in metal detoxification processes. This is in line with a study that combined quantitative susceptibility mapping and imaging transcriptomics and identified that the regions showing the greatest iron deposition in PD were those with a greater expression of genes involved in metal detoxification.^20^

That distinct gene expression and spatial mapping patterns get reflected in the thickness and surface area changes in iRBD is not surprising given that thickness and surface area relate to distinct genetic determinants and developmental trajectories.^32,33^ According to the radial unit hypothesis, cortical thickness corresponds to the number of cells inside the cortical columns, whereas cortical surface area corresponds to the number of ontogenetic columns that populate perpendicularly the cerebral cortex.^53^ Cortical surface area is also subject to tangential expansion due to cellular processes such as synaptogenesis, gliogenesis, and myelination that occur over a longer period than the processes involved in cortical thickness.^54^ Cortical thickness and surface area are differentially affected in PD and iRBD patients and in healthy adults with a higher risk of PD,^15,51,55^ but the biological explanation for this remains unclear. Based on the enrichment patterns identified in this study, it can be hypothesized that unlike thickness, the surface area changes, which related to inflammation, astrocytes and microglia, may not necessarily relate to pathological effects occurring locally. Indeed, microglia and astrocytes are proliferating and circulating cells that can take up alpha-synuclein from the extracellular space, migrate over long distances in the brain, and seed pathology in remote regions.^56^ This is supported by our neighborhood analysis of structural and functional neighbors showing that although the brain architecture did exert a constraining effect on both surface area and thickness, it explained 27-30% of the variance in thickness but only 5-7% of the variance in surface area. In line with that, whereas cortical thickness changes in iRBD could be recreated using a computational spreading model of alpha-synuclein pathology based on gene expression and connectivity, it could not recreate cortical surface area changes,^15^ suggesting that other selective vulnerability or propagation factors explain these changes in iRBD. Based on the findings of the current study, one of these may be the regional density of GABA: in this study, we observed that greater surface area in patients (i.e., the abnormal marker in iRBD) mapped onto regions with lower GABA_A/BZ_ receptor density. In addition to its neurotransmission role, GABA modulates inflammation by regulating the proliferation of immune cells and by decreasing the secretion of cytokines.^57,58^ It may therefore be that changes in cortical surface area occur in regions where there is less potential for GABA to exert its immunomodulatory effects. Although more studies are needed to understand the relationship between surface area and inflammation in the brain of prodromal patients, it nonetheless demonstrates how an approach combining structural MRI, imaging transcriptomics, and comprehensive spatial mapping can be used to generate hypotheses on disease mechanisms in the prodromal phase.

This study has some limitations. First, the gene expression data from the Allen Human Brain Atlas were extracted from post-mortem brains of people aged between 24-57 with different medical histories, causes of death, and post-mortem intervals.^16,59^ Gene expression data were also unavailable for the right hemisphere in 4 of the 6 post-mortem brains.^59^ Future studies should test the gene expression predictors of atrophy when further refined versions of this data atlas become available. Second, the brain atrophy measurements used in this work were derived from T1-weighted images. Post-mortem brain examinations in PD have shown that alpha-synuclein pathology may present first in the axonal processes before being found in cell bodies,^60^ suggesting that assessment of metrics from the white matter may reveal additional information about the dynamics of atrophy occurring in the prodromal phase. Third, the interpretation of findings in this study was limited to patients in the prodromal phase. Future studies should apply a similar processing and analytical framework to atrophy measurements derived from patients with PD and DLB to allow comparability of findings between the prodromal and manifest phases of synucleinopathies. Fourth, only a limited subset of iRBD patients included in this study have so far converted to a manifest synucleinopathy. Once the conversion rate becomes higher, it will be possible to study the atrophy patterns and the gene and connectivity underpinnings in subtypes of iRBD patients based on conversion phenotype.

In sum, the present study shows that mitochondrial and macroautophagy dysfunction underlie the cortical thinning occurring in iRBD and that cortical thinning maps onto specific networks in the brain. With the identification and validation of new imaging-derived markers aimed at predicting prognosis and monitoring disease progression in iRBD, this study highlights the importance of better understanding the mechanisms of the markers we plan to use.

## METHODS

### Participants

A total of 443 participants (182 polysomnography-confirmed iRBD patients and 261 age- and sex-matched healthy controls) with T1-weighted MRI were recruited from five sites: 116 (59 patients) from the Movement Disorders clinic at the Hôpital de la Pitié-Salpêtrière (France), 83 (48 patients) from the Centre for Advanced Research on Sleep Medicine at the Hôpital du Sacré-Cœur de Montréal (Canada), 56 (30 patients) from the ForeFront Parkinson’s Disease Research Clinic at the University of Sydney (Australia), 38 (18 patients) from Aarhus University Hospital (Denmark), and 150 (27 patients) from the Parkinson’s Progression Markers Initiative baseline cohort.^61^ The participants were part of a previous study that assessed brain atrophy in iRBD (see Supp. Table 1 for clinical characterization).^15^ All patients received a polysomnography-proven diagnosis based on the International Classification of Sleep Disorders, third edition.^62^ The absence of concomitant DLB, PD or MSA was confirmed at the neurological evaluation closest in time to the MRI acquisition.^63-65^ Details of the MRI acquisition parameters for each center are reported as Supplementary Information. All participants were part of research protocols approved by local ethics committees, and the current project was approved by the Research Ethics Board of the McGill University Health Centre.

### Quantification of atrophy

Quantification of cortical thickness and surface area in participants was performed previously and region of interest (ROI) results are reported elsewhere.^15^ Briefly, from the 443 T1-weighted images available, 409 scans that passed quality control and underwent surface-based processing with FreeSurfer (version 6.0.0) to generate cortical thickness and surface area maps. All maps were inspected visually by a trained rater (S.R.) based on published guidelines^66,67^ and were excluded if major reconstruction errors (score >2) were found, yielding a final sample of 345 maps from 138 iRBD patients and 207 controls. Cortical thickness and surface area values were next parcellated using the 34-region Desikan-Killiany cortical atlas and *W*-scored to obtain ROI values corrected for the effects of age, sex, and center observed in the matched controls.^22^ Since cortical surface area scales with head size,^68^ surface area ROI values were divided by the FreeSurfer-derived estimated total intracranial volume before *W*-scoring. Cortical thickness and surface area ROI *W*-scores were then *z*-scored and entered in separate PLS regression models to investigate the associations of cortical thinning with transcriptional activity over the whole brain. The primary analyses reported in this article were done using cortical thickness as the predicted variable since we wanted to understand the bases of cortical atrophy in iRBD; the same analyses were however repeated using cortical surface area to better understand the contrast in findings between different cortical metrics.

### Regional gene expression extraction

Regional gene expression was extracted from the same 34 regions as were used for quantifying atrophy. The regional microarray expression data were obtained from 6 post-mortem brains (1 female, mean age (SD, range): 42.5 ± 13.4 (24-57) years) provided by the Allen Human Brain Atlas (https://human.brain-map.org)^59^ and derived using *abagen* (version 0.1.3; https://github.com/rmarkello/abagen).^69^ First, microarray probes were reannotated based on previous data;^16^ probes not matched to a valid Entrez ID were discarded. Next, probes were filtered based on their expression intensity relative to background noise,^70^ such that probes with intensity less than the background in ≥50.0% of samples across donors were discarded. When multiple probes indexed the expression of the same gene, we selected and used the probe with the most consistent pattern of regional variation across donors (i.e., differential stability),^71^ calculated with:

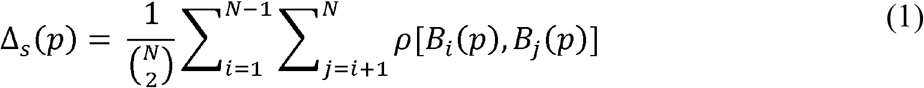

where *ρ* is Spearman’s rank correlation of the expression of a single probe *p* across regions in two donors *B*_*i*_ and *B*_*j*_, and *N* is the total number of donors. Here, regions corresponded to the structural designations provided in the ontology from the Allen Human Brain Atlas.^59^

The MNI coordinates of tissue samples were updated to those generated via non-linear registration using Advanced Normalization Tools (https://github.com/chrisfilo/alleninf). Samples were assigned to brain regions in the provided atlas if their MNI coordinates were within 2 mm of a given parcel. To reduce the potential for misassignment, sample-to-region matching was constrained by hemisphere and gross structural divisions (i.e., cortex, subcortex/brainstem, and cerebellum, such that e.g., a sample in the left cortex could only be assigned to an atlas parcel in the left cortex).^16^ All tissue samples not assigned to a brain region in the provided atlas were discarded.

Inter-subject variation was addressed by normalizing tissue sample expression values across genes using a robust sigmoid function,^72^ as follows:

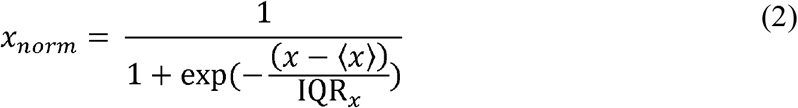

where ⟨*x* ⟩ is the median and IQR_*X*_ is the normalized interquartile range of the expression of a single tissue sample across genes. Normalized expression values were then rescaled to the unit interval:

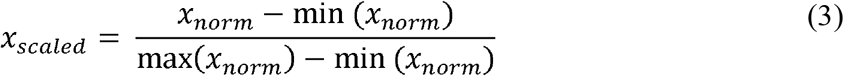

Gene expression values were then normalized across tissue samples using an identical procedure. Samples assigned to the same brain region were averaged separately for each donor and then across donors, yielding a regional expression matrix. Since 4 of the 6 post-mortem brains did not have available gene expression in the right hemisphere, analyses involving gene expression were only performed in regions from the left hemisphere. The resulting gene expression ROI values were *z*-scored and entered as predictors in PLS regression.

### PLS regression

PLS regression was used to identify the pattern of gene expression that predicted cortical thickness *W*-scores in iRBD. PLS regression is a multivariate data reduction technique that performs a dual decomposition of two matrices **X** and **Y** to derive components from **X** (34 regions x 15,633 genes) that account for the maximal amount of covariance explained by **Y** (34 cortical *W*-scores). To test the null hypothesis that the derived components did not predict more variance in morphology than chance, the empirical variance was tested against the variance observed in 10,000 null models in which atrophy was randomly permuted between regions (random null models). Since the brain is characterized by a high level of spatial autocorrelation between regions,^73^ to demonstrate that the gene-atrophy associations were not solely due to the variance attributable to lower-order spatial gradients, the empirical variance was also compared to 10,000 spatially-constrained null models in which regions were shuffled using a spherical reassignment procedure that preserved spatial autocorrelation between regions (spatial null models).^73^ The coordinates inside each parcel were averaged using the *netneurotools* toolbox (https://github.com/netneurolab/netneurotools) and the parcel centroids corresponded to the vertices on the sphere with the lowest Euclidean distance. A PLS component was considered significant when less than 5% of null models explained more variance than the original atrophy vector.

To identify the genes that contributed the most to the significant components associated with cortical changes, a bootstrapping resampling procedure was performed by shuffling randomly the rows of **X** and **Y** and by repeating the PLS regression with the shuffled matrices; this was repeated 5,000 times to generate the null distribution and the standard errors for each gene expression weight. Bootstrap ratios were calculated as the ratio between the weight of each gene expression and its bootstrap-estimated standard error and interpreted as a *z*-score.^74^ The gene lists were then ranked from the highest to the lowest scores based on the bootstrap ratios and used as inputs for GSEA. The PLS analysis was also repeated for predicting the pattern of cortical surface area in iRBD instead of cortical thickness.

### Gene enrichment analyses

To examine the translational relevance of transcriptomic correlates of atrophy in iRBD, WebGestalt 2019 (http://www.webgestalt.org)^75^ was used to perform GSEA and identify the biological process, cellular component, and disease-related gene terms enriched in the genes predicting cortical thickness *W*-scores in iRBD. GSEA assesses whether the genes located at the top or bottom of a ranked gene list, in this case, derived from the bootstrapping resampling procedure, occurred more frequently than expected by chance.^76^ GSEA was performed separately for patterns of thickness *W*-scores. The Gene Ontology knowledge base (April 2019, http://geneontology.org) was used for biological process and cellular component gene terms, whereas the DisGeNET (version 5.0, May 2017, https://www.disgenet.org), OMIM (https://www.omim.org), and GLAD4U (November 2018, http://glad4u.zhang-lab.org/index.php) knowledge bases were used separately for disease-related gene terms. The minimal and maximal number of genes for enrichment was set to 3 and 2,000, respectively. Statistical correction for multiple testing was performed by running 1,000 random permutations and adjusting *P*-values with the false discovery rate (FDR) method; for interpretability, only the top ten most significant terms retrieved on the positive and negative ends were interpreted.

We also performed over-representation analysis in WebGestalt 2019 to investigate whether the genes most reliably associated with thickness *W*-scores were enriched for gene terms related to specific biological processes, cellular components, and diseases compared to the complete gene set. The target list was composed of the genes with a bootstrap ratio weight ±5.0 (corresponding to *z*-scores less than *p*<0.0001), whereas the background list was composed of all the genes extracted from the Allen Human Brain Atlas. To ensure that the enrichment patterns were not due to the choice of a particular gene ontology platform, we repeated the over-representation analyses using GOrilla (http://cbl-gorilla.cs.technion.ac.il)^77^ to assess the biological process and cellular component gene terms enriched in the target lists (i.e., negatively or positively weighted genes with bootstrap ratios ±5.0 predicting cortical thickness *W*-scores in iRBD) compared to the background list (i.e., all genes extracted from the Allen Human Brain Atlas).^59^ These enrichment analyses was also repeated on the target lists resulting from the PLS analysis applied to cortical surface area.

### Virtual histology

We next used a virtual histology approach to investigate whether the genes associated with specific cell types in the brain were more pronounced in the genes predicting cortical thickness *W*-scores in iRBD. Using single-cell RNA sequencing post-mortem studies performed on human cortical samples,^17,24^ the regional expression of the genes associated with astrocytes, endothelial cells, microglia, excitatory neurons, inhibitory neurons, oligodendrocytes, and oligodendrocyte precursor cells were extracted in *abagen* for the same 34 regions as used for quantifying cortical morphology and averaged across genes to generate an average regional gene expression for each of the 7 cell types.^28^ Spearman’s correlation coefficients were computed between each cell type’s average regional gene expression and the regional cortical thickness *W*-scores. An association was considered significant when below the Bonferroni-corrected threshold of *p*<0.0071 (7 correlations) and tested against 10,000 null models with and without preservation of the spatial autocorrelation between brain regions (spatial and random null models, respectively). The same analysis was also repeated using cortical surface area instead of cortical thickness.

### Structural and functional neighborhood analysis

According to the prion-like spread hypothesis, pathology is expected to spread between cells following the constraints imposed by the brain’s architecture (connectivity).^48^ Here, we tested whether structural and functional connectivity constrained the cortical changes observed in iRBD by assessing whether the cortical thickness *W*-scores of each region were associated with the average cortical thickness *W*-scores measured in the structurally or functionally connected neighbors of each region. Unlike analyses involving gene expression, which were restricted to the left hemisphere, these network analyses were performed using both hemispheres. To identify the structural and functional neighbors of each region, the diffusion-weighted and resting-state functional MRI data from 70 healthy participants were used to generate individual structural and functional connectivity maps. These data were processed previously^78^ and used in several studies assessing the impact of connectivity on atrophy.^17,79-81^ After exclusion of 4 connectivity maps with aberrant scores (3 structural, 1 functional), the individual structural matrices were transformed into a group-consensus structural connectivity matrix that preserved the brain’s edge length distribution;^82^ self-connections were converted to zero.

To assess the impact of structural connectivity on cortical thickness in iRBD, the structural neighborhood atrophy *D*_*i*_ was quantified by averaging the atrophy observed in the total number *N* of nodes *j* that had a structural connection with each region *i* (i.e., structural neighbors) using the following formula:

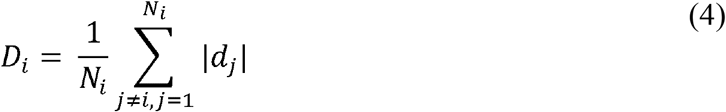

To assess the impact of functional connectivity on cortical atrophy in iRBD, the functional neighborhood atrophy was calculated by averaging the atrophy observed in all the structural neighbors *j* weighted by the strength of the functional connection between region *i* and neighbor *j* (*FC*_*ij*_), using the following formula:

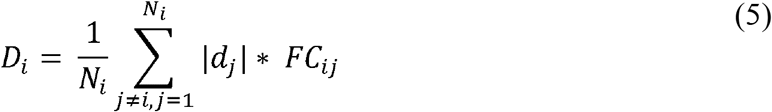

Spearman’s correlation coefficients were then calculated between the regional cortical thickness *W*-scores in iRBD, and the structural and functional neighborhood atrophy quantified in each region. The empirical correlation coefficients were next tested against the correlations observed in sets of 10,000 randomized null models that were randomly shuffled with and without preserving the brain’s spatial autocorrelation (spatial and random null models). The analyses were repeated for cortical surface area.

### Spatial mapping of cortical changes to brain systems

We next performed a comprehensive mapping study of the cortical changes in iRBD by assessing whether the cortical thickness *W*-scores were significantly more pronounced in specific brain systems, namely neurotransmitter density, intrinsic functional networks, cytoarchitectonic classes, and spatial correlates of cognitive functions. For neurotransmitter density, the previously curated regional density maps of 18 receptors, transporters, and receptor binding sites associated with dopamine (D1, D2, DAT), serotonin (5-HT_1A_, 5-HT_1B_, 5-HT_2A_, 5-HT_4_, 5-HT_6_, 5-HTT), noradrenaline (NET), acetylcholine (α4β2, M_1_, VAChT), GABA (GABA_A/BZ_), glutamate (mGluR5), histamine (H_3_), endocannabinoids (CB_1_), and opioids (μ), were generated from the PET images of more than 1,200 healthy individuals (see Supplementary Information 2 for the articles chosen to construct each map).^28^ These density maps were parcellated into the same 68 regions used for deriving atrophy, and each map was *z*-scored separately. When several maps were available for a tracer, a weighted average map was generated based on the number of subjects used for each map. Spearman’s correlation coefficients were used to test the association between regional cortical thickness *W*-scores and the regional density of each of the 18 receptors, transporters, and binding sites. Correlations surviving the Bonferroni-corrected threshold for each set of associations (*p*<0.0028) were tested against sets of 10,000 randomized null models with and without preservation of the brain’s spatial autocorrelation (spatial and random null models).

We next investigated whether cortical thickness *W*-scores in iRBD were more pronounced in regions with a specific functional time course and laminar organization, using respectively the Yeo parcellation^29,83^ and the extended version of the von Economo and Koskinas atlas.^31,83,84^ The Yeo parcellation scheme attributes every brain region to one of 7 resting-state networks (i.e., visual, sensorimotor, dorsal attention, ventral attention, limbic, frontoparietal, and default-mode networks), whereas the cytoarchitectonic classes from the von Economo and Koskinas atlas attribute every region to one of 7 cortical lamination classes (i.e., primary motor cortex, association cortices 1 and 2, primary and secondary sensory areas (dysgranular cortex), primary sensory cortex (agranular cortex), limbic regions, and insular cortex). For each network and class, the cortical thickness *W*-scores were averaged and tested against sets of 10,000 randomized null models with and without the preservation of the brain’s spatial autocorrelation (spatial and random null models).

We also tested the correspondence between the patterns of cortical thickness *W*-scores in iRBD and the spatial correlates associated with several cognitive processes. A total of 123 cognitive processes from the Cognitive Atlas (https://www.cognitiveatlas.org)^28,85^ were selected and their activation maps were obtained from the meta-analytic activation maps available as part of the Neurosynth database (http://www.neurosynth.org).^86^ Every regional value represented the probability of each of the 68 cortical regions used for deriving atrophy in iRBD to be activated during the functional task associated with the cognitive process. Spearman’s correlations were performed to test the association between cortical thickness W-scores and the regional functional activation pattern of each of the 123 cognitive processes. Correlations surviving the Bonferroni-corrected threshold (p<0.00041) were tested against sets of 10,000 randomized null models with and without preservation of the brain’s spatial autocorrelation (spatial and random null models). This spatial mapping approach was repeated for cortical surface area in iRBD.

## Data Availability

The regional cortical thickness and surface area values are available at https://github.com/srahayel/SIR-RBD. Codes used for performing the genetic and connectivity analyses are available publicly from the cited references.

## DATA AVAILABILITY

The regional cortical thickness and surface area values are available at https://github.com/srahayel/SIR-RBD.

## CODE AVAILABILITY

Codes used for performing the genetic and connectivity analyses are available publicly from the cited references.

## ACKNOWLEDGMENTS

Shady Rahayel reports a scholarship from the Canadian Institutes of Health Research.

The work performed in Paris was funded by grants from the Programme d’investissements d’avenir (ANR-10-IAIHU-06), the Paris Institute of Neurosciences – IHU (IAIHU-06), the Agence Nationale de la Recherche (ANR-11-INBS-0006), Électricité de France (Fondation d’Entreprise EDF), Biogen Inc., the Fondation Thérèse et René Planiol, the Fonds Saint-Michel; by unrestricted support for research on Parkinson’s disease from Energipole (M. Mallart) and Société Française de Médecine Esthétique (M. Legrand); and by a grant from the Institut de France to Isabelle Arnulf (for the Alice Study).

The work performed in Montreal was supported by the Canadian Institutes of Health Research, the Fonds de recherche du Québec–Santé, and the W. Garfield Weston Foundation. Jean-François Gagnon holds a Canada Research Chair in Cognitive Decline in Pathological Aging and reports grants from the Fonds de recherche du Québec – Santé, the Canadian Institutes of Health Research, the W. Garfield Weston Foundation, the Michael J. Fox Foundation for Parkinson’s Research, and the National Institutes of Health. Ronald B. Postuma reports grants and personal fees from the Fonds de recherche du Québec–Santé, the Canadian Institutes of Health Research, the Parkinson Society of Canada, the W. Garfield Weston Foundation, the Michael J. Fox Foundation for Parkinson’s Research, the R. Howard Webster Foundation, and the National Institutes of Health. Ziv Gan-Or reports funding from the Michael J. Fox Foundation, Canadian Consortium for Neurodegeneration in Aging, Fonds de recherche du Québec–Santé, and ‘Healthy Brains, Healthy lives’ initiative. Ziv Gan-Or is supported by the Fonds de recherche du Québec– Santé Chercheur-Boursier award and is a William Dawson Scholar. This work was also funded by awards from the Canadian Institutes of Health Research Foundation Scheme and the ‘Healthy Brain, Healthy Lives’ initiative to Alain Dagher.

The work performed in Sydney was supported by a Dementia Team Grant from the National Health and Medical Research Council (#1095127). Simon Lewis is supported by a Leadership Fellowship from the National Health and Medical Research Council (#1195830). Elie Matar reports funding from the National Health and Medical Research Council (#2008565). Kaylena Ehgoetz Martens reports funding from Parkinson Canada, Parkinson’s Movement Disorder Foundation, University of Waterloo International Research Partnership grant, and Natural Science and Engineering Research Council of Canada.

The work performed in Aarhus was supported by funding from the Lundbeck Foundation, Parkinsonforeningen (The Danish Parkinson Association), and the Jascha Foundation.

## AUTHOR CONTRIBUTIONS

Conceptualization: S.R., Z.G.O., A.D.; methodology: S.R., C.T., A.V., B.M., Z.G.O., A.D.; formal analysis: S.R., C.T., A.V.; clinical investigation: S.Leh., I.A., M.V., J.C.C., J.F.G., R.B.P., J.M., S.Lew., E.M., K.E.M., P.B., K.K., A.K.H., O.M., Z.G.O., A.D.; writing-original draft: S.R.; writing-review and editing: all authors; visualization: S.R.; supervision: Z.G.O., A.D.; funding acquisition: B.M., S.Leh., I.A., M.V., J.C.C., J.F.G., R.B.P., J.M., S.Lew., E.M., K.E.M., P.B., K.K., A.K.H., O.M., Z.G.O., A.D.

## REFERENCES

1. Postuma RB, Iranzo A, Hu M, et al. Risk and predictors of dementia and parkinsonism in idiopathic REM sleep behaviour disorder: a multicentre study. Brain. Mar 1 2019;142(3):744–759. doi:10.1093/brain/awz030

2. Hogl B, Stefani A, Videnovic A. Idiopathic REM sleep behaviour disorder and neurodegeneration - an update. Nat Rev Neurol. Jan 2018;14(1):40–55. doi:10.1038/nrneurol.2017.157

3. Bourgouin PA, Rahayel S, Gaubert M, et al. Neuroimaging of Rapid Eye Movement Sleep Behavior Disorder. Int Rev Neurobiol. 2019;144:185–210. doi:10.1016/bs.irn.2018.10.006

4. Campabadal A, Segura B, Junque C, Iranzo A. Structural and functional magnetic resonance imaging in isolated REM sleep behavior disorder: A systematic review of studies using neuroimaging software. Sleep Med Rev. Apr 22 2021;59:101495. doi:10.1016/j.smrv.2021.101495

5. Rahayel S, Postuma RB, Montplaisir J, et al. Abnormal Gray Matter Shape, Thickness, and Volume in the Motor Cortico-Subcortical Loop in Idiopathic Rapid Eye Movement Sleep Behavior Disorder: Association with Clinical and Motor Features. Cereb Cortex. Feb 1 2018;28(2):658–671. doi:10.1093/cercor/bhx137

6. Rahayel S, Postuma RB, Montplaisir J, et al. Cortical and subcortical gray matter bases of cognitive deficits in REM sleep behavior disorder. Neurology. May 15 2018;90(20):e1759–e1770. doi:10.1212/WNL.0000000000005523

7. Campabadal A, Segura B, Junque C, et al. Comparing the accuracy and neuroanatomical correlates of the UPSIT-40 and the Sniffin’ Sticks test in REM sleep behavior disorder. Parkinsonism Relat Disord. Aug 2019;65:197–202. doi:10.1016/j.parkreldis.2019.06.013

8. Pereira JB, Weintraub D, Chahine L, Aarsland D, Hansson O, Westman E. Cortical thinning in patients with REM sleep behavior disorder is associated with clinical progression. NPJ Parkinsons Dis. 2019;5:7. doi:10.1038/s41531-019-0079-3

9. Rahayel S, Postuma RB, Montplaisir J, et al. A Prodromal Brain-Clinical Pattern of Cognition in Synucleinopathies. Ann Neurol. Feb 2021;89(2):341–357. doi:10.1002/ana.25962

10. Kordower JH, Chu Y, Hauser RA, Freeman TB, Olanow CW. Lewy body-like pathology in long-term embryonic nigral transplants in Parkinson’s disease. Nat Med. May 2008;14(5):504–6. doi:10.1038/nm1747

11. Gonzalez-Rodriguez P, Zampese E, Surmeier DJ. Selective neuronal vulnerability in Parkinson’s disease. Prog Brain Res. 2020;252:61–89. doi:10.1016/bs.pbr.2020.02.005

12. Luk KC, Kehm V, Carroll J, et al. Pathological alpha-synuclein transmission initiates Parkinson-like neurodegeneration in nontransgenic mice. Science. Nov 16 2012;338(6109):949–53. doi:10.1126/science.1227157

13. Zheng YQ, Zhang Y, Yau Y, et al. Local vulnerability and global connectivity jointly shape neurodegenerative disease propagation. PLoS Biol. Nov 2019;17(11):e3000495. doi:10.1371/journal.pbio.3000495

14. Rahayel S, Misic B, Zheng YQ, et al. Differentially targeted seeding reveals unique pathological alpha-synuclein propagation patterns. Brain. Jun 3 2022;145(5):1743–1756. doi:10.1093/brain/awab440

15. Rahayel S, Tremblay C, Vo A, et al. Brain atrophy in prodromal synucleinopathy is shaped by structural connectivity and gene expression. Brain. May 20 2022;doi:10.1093/brain/awac187

16. Arnatkeviciute A, Fulcher BD, Fornito A. A practical guide to linking brain-wide gene expression and neuroimaging data. Neuroimage. Apr 1 2019;189:353–367. doi:10.1016/j.neuroimage.2019.01.011

17. Tremblay C, Rahayel S, Vo A, et al. Brain atrophy progression in Parkinson’s disease is shaped by connectivity and local vulnerability. Brain Commun. 2021;3(4):fcab269. doi:10.1093/braincomms/fcab269

18. Freeze B, Pandya S, Zeighami Y, Raj A. Regional transcriptional architecture of Parkinson’s disease pathogenesis and network spread. Brain. Oct 1 2019;142(10):3072–3085. doi:10.1093/brain/awz223

19. Maia PD, Pandya S, Freeze B, et al. Origins of atrophy in Parkinson linked to early onset and local transcription patterns. Brain Commun. 2020;2(2):fcaa065. doi:10.1093/braincomms/fcaa065

20. Thomas GEC, Zarkali A, Ryten M, et al. Regional brain iron and gene expression provide insights into neurodegeneration in Parkinson’s disease. Brain. Jul 28 2021;144(6):1787–1798. doi:10.1093/brain/awab084

21. Brown JA, Deng J, Neuhaus J, et al. Patient-Tailored, Connectivity-Based Forecasts of Spreading Brain Atrophy. Neuron. Dec 4 2019;104(5):856–868 e5. doi:10.1016/j.neuron.2019.08.037

22. Tremblay C, Abbasi N, Zeighami Y, et al. Sex effects on brain structure in de novo Parkinson’s disease: a multimodal neuroimaging study. Brain. Oct 1 2020;143(10):3052–3066. doi:10.1093/brain/awaa234

23. La Joie R, Perrotin A, Barre L, et al. Region-specific hierarchy between atrophy, hypometabolism, and beta-amyloid (Abeta) load in Alzheimer’s disease dementia. J Neurosci. Nov 14 2012;32(46):16265–73. doi:10.1523/JNEUROSCI.2170-12.2012

24. Seidlitz J, Nadig A, Liu S, et al. Transcriptomic and cellular decoding of regional brain vulnerability to neurogenetic disorders. Nat Commun. Jul 3 2020;11(1):3358. doi:10.1038/s41467-020-17051-5

25. Rey NL, George S, Steiner JA, et al. Spread of aggregates after olfactory bulb injection of alpha-synuclein fibrils is associated with early neuronal loss and is reduced long term. Acta Neuropathol. Jan 2018;135(1):65–83. doi:10.1007/s00401-017-1792-9

26. Recasens A, Dehay B, Bove J, et al. Lewy body extracts from Parkinson disease brains trigger alpha-synuclein pathology and neurodegeneration in mice and monkeys. Ann Neurol. Mar 2014;75(3):351–62. doi:10.1002/ana.24066

27. Li JY, Englund E, Holton JL, et al. Lewy bodies in grafted neurons in subjects with Parkinson’s disease suggest host-to-graft disease propagation. Nat Med. May 2008;14(5):501–3. doi:10.1038/nm1746

28. Hansen JY, Shafiei G, Markello RD, et al. Mapping neurotransmitter systems to the structural and functional organization of the human neocortex. bioRxiv. 2022:2021.10.28.466336. doi:10.1101/2021.10.28.466336

29. Yeo BT, Krienen FM, Sepulcre J, et al. The organization of the human cerebral cortex estimated by intrinsic functional connectivity. J Neurophysiol. Sep 2011;106(3):1125–65. doi:10.1152/jn.00338.2011

30. von Economo C, Koskinas GN. Die Cytoarchitektonik der Hirnrinde des erwachsenen Menschen. (The Cyto-Architectonics of the Cerebral Cortex of Adult Man.). 1925.

31. Vertes PE, Rittman T, Whitaker KJ, et al. Gene transcription profiles associated with inter-modular hubs and connection distance in human functional magnetic resonance imaging networks. Philos Trans R Soc Lond B Biol Sci. Oct 5 2016;371(1705)doi:10.1098/rstb.2015.0362

32. Hogstrom LJ, Westlye LT, Walhovd KB, Fjell AM. The structure of the cerebral cortex across adult life: age-related patterns of surface area, thickness, and gyrification. Cereb Cortex. Nov 2013;23(11):2521–30. doi:10.1093/cercor/bhs231

33. Grasby KL, Jahanshad N, Painter JN, et al. The genetic architecture of the human cerebral cortex. Science. Mar 20 2020;367(6484)doi:10.1126/science.aay6690

34. Panizzon MS, Fennema-Notestine C, Eyler LT, et al. Distinct genetic influences on cortical surface area and cortical thickness. Cereb Cortex. Nov 2009;19(11):2728–35. doi:10.1093/cercor/bhp026

35. Rahayel S, Montplaisir J, Monchi O, et al. Patterns of cortical thinning in idiopathic rapid eye movement sleep behavior disorder. Mov Disord. Apr 15 2015;30(5):680–7. doi:10.1002/mds.25820

36. Holtbernd F, Romanzetti S, Oertel WH, et al. Convergent patterns of structural brain changes in rapid eye movement sleep behavior disorder and Parkinson’s disease on behalf of the German rapid eye movement sleep behavior disorder study group. Sleep. Mar 12 2021;44(3)doi:10.1093/sleep/zsaa199

37. Campabadal A, Segura B, Junque C, et al. Cortical Gray Matter and Hippocampal Atrophy in Idiopathic Rapid Eye Movement Sleep Behavior Disorder. Front Neurol. 2019;10:312. doi:10.3389/fneur.2019.00312

38. Tansey MG, Wallings RL, Houser MC, Herrick MK, Keating CE, Joers V. Inflammation and immune dysfunction in Parkinson disease. Nat Rev Immunol. Mar 4 2022;doi:10.1038/s41577-022-00684-6

39. Haelterman NA, Yoon WH, Sandoval H, Jaiswal M, Shulman JM, Bellen HJ. A mitocentric view of Parkinson’s disease. Annu Rev Neurosci. 2014;37:137–59. doi:10.1146/annurev-neuro-071013-014317

40. Gonzalez-Rodriguez P, Zampese E, Stout KA, et al. Disruption of mitochondrial complex I induces progressive parkinsonism. Nature. Nov 2021;599(7886):650–656. doi:10.1038/s41586-021-04059-0

41. Subrahmanian N, LaVoie MJ. Is there a special relationship between complex I activity and nigral neuronal loss in Parkinson’s disease? A critical reappraisal. Brain Res. Sep 15 2021;1767:147434. doi:10.1016/j.brainres.2021.147434

42. Parker WD, Jr., Parks JK, Swerdlow RH. Complex I deficiency in Parkinson’s disease frontal cortex. Brain Res. Jan 16 2008;1189:215–8. doi:10.1016/j.brainres.2007.10.061

43. Keeney PM, Xie J, Capaldi RA, Bennett JP, Jr. Parkinson’s disease brain mitochondrial complex I has oxidatively damaged subunits and is functionally impaired and misassembled. J Neurosci. May 10 2006;26(19):5256–64. doi:10.1523/JNEUROSCI.0984-06.2006

44. Griffey CJ, Yamamoto A. Macroautophagy in CNS health and disease. Nat Rev Neurosci. May 3 2022;doi:10.1038/s41583-022-00588-3

45. Merino-Galan L, Jimenez-Urbieta H, Zamarbide M, et al. Striatal synaptic bioenergetic and autophagic decline in premotor experimental parkinsonism. Brain. Mar 4 2022;doi:10.1093/brain/awac087

46. Grassi D, Howard S, Zhou M, et al. Identification of a highly neurotoxic alpha-synuclein species inducing mitochondrial damage and mitophagy in Parkinson’s disease. Proc Natl Acad Sci U S A. Mar 13 2018;115(11):E2634–E2643. doi:10.1073/pnas.1713849115

47. Volpicelli-Daley LA, Luk KC, Patel TP, et al. Exogenous alpha-synuclein fibrils induce Lewy body pathology leading to synaptic dysfunction and neuron death. Neuron. Oct 6 2011;72(1):57–71. doi:10.1016/j.neuron.2011.08.033

48. Peng C, Trojanowski JQ, Lee VM. Protein transmission in neurodegenerative disease. Nat Rev Neurol. Apr 2020;16(4):199–212. doi:10.1038/s41582-020-0333-7

49. Tremblay C, Rahayel S, Vo A, et al. Brain atrophy progression in Parkinson’s disease is shaped by connectivity and local vulnerability. medRxiv. 2021:2021.06.08.21258321. doi:10.1101/2021.06.08.21258321

50. Fjell AM, Westlye LT, Grydeland H, et al. Accelerating cortical thinning: unique to dementia or universal in aging? Cereb Cortex. Apr 2014;24(4):919–34. doi:10.1093/cercor/bhs379

51. Abbasi N, Tremblay C, Rajimehr R, et al. Neuroanatomical correlates of polygenic risk for Parkinson’s Disease. medRxiv. 2022:2022.01.17.22269262. doi:10.1101/2022.01.17.22269262

52. Bottigliengo D, Foco L, Seibler P, Klein C, Konig IR, Del Greco MF. A Mendelian randomization study investigating the causal role of inflammation on Parkinson’s disease. Brain. Jun 3 2022;doi:10.1093/brain/awac193

53. Rakic P. Radial unit hypothesis of neocortical expansion. Novartis Found Symp. 2000;228:30-42; discussion 42-52. doi:10.1002/0470846631.ch3

54. Cafiero R, Brauer J, Anwander A, Friederici AD. The Concurrence of Cortical Surface Area Expansion and White Matter Myelination in Human Brain Development. Cereb Cortex. Feb 1 2019;29(2):827–837. doi:10.1093/cercor/bhy277

55. Laansma MA, Bright JK, Al-Bachari S, et al. International Multicenter Analysis of Brain Structure Across Clinical Stages of Parkinson’s Disease. Mov Disord. Nov 2021;36(11):2583–2594. doi:10.1002/mds.28706

56. Steiner JA, Quansah E, Brundin P. The concept of alpha-synuclein as a prion-like protein: ten years after. Cell Tissue Res. Jul 2018;373(1):161–173. doi:10.1007/s00441-018-2814-1

57. Jin Z, Mendu SK, Birnir B. GABA is an effective immunomodulatory molecule. Amino Acids. Jul 2013;45(1):87–94. doi:10.1007/s00726-011-1193-7

58. Crowley T, Cryan JF, Downer EJ, O’Leary OF. Inhibiting neuroinflammation: The role and therapeutic potential of GABA in neuro-immune interactions. Brain Behav Immun. May 2016;54:260–277. doi:10.1016/j.bbi.2016.02.001

59. Hawrylycz MJ, Lein ES, Guillozet-Bongaarts AL, et al. An anatomically comprehensive atlas of the adult human brain transcriptome. Nature. Sep 20 2012;489(7416):391–399. doi:10.1038/nature11405

60. Braak H, Sandmann-Keil D, Gai W, Braak E. Extensive axonal Lewy neurites in Parkinson’s disease: a novel pathological feature revealed by alpha-synuclein immunocytochemistry. Neurosci Lett. Apr 9 1999;265(1):67–9. doi:10.1016/s0304-3940(99)00208-6

61. Marek K, Chowdhury S, Siderowf A, et al. The Parkinson’s progression markers initiative (PPMI) - establishing a PD biomarker cohort. Ann Clin Transl Neurol. Dec 2018;5(12):1460–1477. doi:10.1002/acn3.644

62. American Academy of Sleep Medicine. The International Classification of Sleep Disorders — Third Edition (ICSD-3). 3rd ed. American Academy of Sleep Medicine; 2014.

63. Postuma RB, Berg D, Stern M, et al. MDS clinical diagnostic criteria for Parkinson’s disease. Mov Disord. Oct 2015;30(12):1591–601. doi:10.1002/mds.26424

64. McKeith IG, Boeve BF, Dickson DW, et al. Diagnosis and management of dementia with Lewy bodies: Fourth consensus report of the DLB Consortium. Neurology. Jul 4 2017;89(1):88–100. doi:10.1212/WNL.0000000000004058

65. Gilman S, Wenning GK, Low PA, et al. Second consensus statement on the diagnosis of multiple system atrophy. Neurology. Aug 26 2008;71(9):670–6. doi:10.1212/01.wnl.0000324625.00404.15

66. Klapwijk ET, van de Kamp F, van der Meulen M, Peters S, Wierenga LM. Qoala-T: A supervised-learning tool for quality control of FreeSurfer segmented MRI data. Neuroimage. Apr 1 2019;189:116–129. doi:10.1016/j.neuroimage.2019.01.014

67. Monereo-Sanchez J, de Jong JJA, Drenthen GS, et al. Quality control strategies for brain MRI segmentation and parcellation: Practical approaches and recommendations - insights from the Maastricht study. Neuroimage. Aug 15 2021;237:118174. doi:10.1016/j.neuroimage.2021.118174

68. Barnes J, Ridgway GR, Bartlett J, et al. Head size, age and gender adjustment in MRI studies: a necessary nuisance? Neuroimage. Dec 2010;53(4):1244–55. doi:10.1016/j.neuroimage.2010.06.025

69. Markello RD, Arnatkevičiūtė A, Poline J-B, Fulcher BD, Fornito A, Misic B. Standardizing workflows in imaging transcriptomics with the abagen toolbox. bioRxiv. 2021:2021.07.08.451635. doi:10.1101/2021.07.08.451635

70. Quackenbush J. Microarray data normalization and transformation. Nat Genet. Dec 2002;32 Suppl:496–501. doi:10.1038/ng1032

71. Hawrylycz M, Miller JA, Menon V, et al. Canonical genetic signatures of the adult human brain. Nat Neurosci. Dec 2015;18(12):1832–44. doi:10.1038/nn.4171

72. Fulcher BD, Little MA, Jones NS. Highly comparative time-series analysis: the empirical structure of time series and their methods. J R Soc Interface. Jun 6 2013;10(83):20130048. doi:10.1098/rsif.2013.0048

73. Vasa F, Misic B. Null models in network neuroscience. Nat Rev Neurosci. Aug 2022;23(8):493–504. doi:10.1038/s41583-022-00601-9

74. Efron B, Tibshirani R. Bootstrap methods for standard errors, confidence intervals, and other measures of statistical accuracy. Stat Sci. 1986;1:54–77.

75. Liao Y, Wang J, Jaehnig EJ, Shi Z, Zhang B. WebGestalt 2019: gene set analysis toolkit with revamped UIs and APIs. Nucleic Acids Res. Jul 2 2019;47(W1):W199–W205. doi:10.1093/nar/gkz401

76. Subramanian A, Tamayo P, Mootha VK, et al. Gene set enrichment analysis: a knowledge-based approach for interpreting genome-wide expression profiles. Proc Natl Acad Sci U S A. Oct 25 2005;102(43):15545–50. doi:10.1073/pnas.0506580102

77. Eden E, Navon R, Steinfeld I, Lipson D, Yakhini Z. GOrilla: a tool for discovery and visualization of enriched GO terms in ranked gene lists. BMC Bioinformatics. Feb 3 2009;10:48. doi:10.1186/1471-2105-10-48

78. Cammoun L, Gigandet X, Meskaldji D, et al. Mapping the human connectome at multiple scales with diffusion spectrum MRI. J Neurosci Methods. Jan 30 2012;203(2):386–97. doi:10.1016/j.jneumeth.2011.09.031

79. Shafiei G, Markello RD, Makowski C, et al. Spatial Patterning of Tissue Volume Loss in Schizophrenia Reflects Brain Network Architecture. Biol Psychiatry. Apr 15 2020;87(8):727–735. doi:10.1016/j.biopsych.2019.09.031

80. Hansen JY, Shafiei G, Vogel JW, et al. Molecular and connectomic vulnerability shape cross-disorder cortical abnormalities. bioRxiv. 2022:2022.01.21.476409. doi:10.1101/2022.01.21.476409

81. Shafiei G, Bazinet V, Dadar M, et al. Network structure and transcriptomic vulnerability shape atrophy in frontotemporal dementia. Brain. Feb 21 2022;doi:10.1093/brain/awac069

82. Betzel RF, Griffa A, Hagmann P, Misic B. Distance-dependent consensus thresholds for generating group-representative structural brain networks. Netw Neurosci. 2019;3(2):475–496. doi:10.1162/netn_a_00075

83. Vazquez-Rodriguez B, Suarez LE, Markello RD, et al. Gradients of structure-function tethering across neocortex. Proc Natl Acad Sci U S A. Oct 15 2019;116(42):21219–21227. doi:10.1073/pnas.1903403116

84. Scholtens LH, de Reus MA, de Lange SC, Schmidt R, van den Heuvel MP. An MRI Von Economo - Koskinas atlas. Neuroimage. Apr 15 2018;170:249–256. doi:10.1016/j.neuroimage.2016.12.069

85. Poldrack RA, Kittur A, Kalar D, et al. The cognitive atlas: toward a knowledge foundation for cognitive neuroscience. Front Neuroinform. 2011;5:17. doi:10.3389/fninf.2011.00017

86. Yarkoni T, Poldrack RA, Nichols TE, Van Essen DC, Wager TD. Large-scale automated synthesis of human functional neuroimaging data. Nat Methods. Jun 26 2011;8(8):665–70. doi:10.1038/nmeth.1635

