## Supplemental Material for "Mitochondrial dysfunction underlies cortical atrophy in prodromal synucleinopathies"

**Supplementary Information 1.**

Details of T1-weighted MRI acquisition in the different centers.

The Montreal cohort underwent T1-weighted imaging with a 3T Siemens TIM Trio scanner with a 12-channel head coil, MPRAGE sequence: TR: 2300 ms, TE: 2.91 ms, flip angle: 9°, and voxel size: 1 mm³ isotropic.

The Paris cohort underwent T1-weighted imaging with a 3T Siemens TIM Trio scanner with a 12-channel head coil, MPRAGE sequence: TR: 2300 ms, TE: 4.18 ms, TI: 900 ms, flip angle: 9°, and voxel size: 1 mm³ isotropic; or a 3T PRISMA Fit scanner with a 64-channel head coil, MP2RAGE sequence: TR: 5,000 ms, TE: 2.98 ms, TI: 700 and 2500 ms, flip angle: 4° and 5°, GRAPPA: 3, and voxel size: 1 mm³ isotropic.

The Sydney cohort was imaged with a GE Discovery MR750 3T scanner with an 8-channel head coil, BRAVO sequence: TR: 5800 ms, TE: 2.6 ms, flip angle: 12°, and voxel size: 1 mm³ isotropic.

The Aarhus cohort was imaged with a 3T Siemens MAGNETOM Skyra scanner with a 32-channel head coil, MPRAGE sequence: TR: 2,420 ms, TE: 3.7 ms, TI: 960 ms, flip angle: 9°, and voxel size: 1 mm³ isotropic.

The T1-weighted images from the Parkinson’s Progression Markers Initiative cohort are described elsewhere (Marek et al., 2018).

**Supplementary Information 2.**

Supplementary references of the articles used for constructing each receptor, transporter, and receptor binding site map (from (Hansen et al., 2022).

Dopamine:

- D1: Kaller et al. (2017)
- D2: (Sandiego et al., 2015; Smith et al., 2019)
- DAT: (Dukart et al., 2018)

Serotonin:

- 5-HT_1A_: (Savli et al., 2012)
- 5-HT_1B_: (Gallezot et al., 2010; Savli et al., 2012)
- 5-HT_2A_: (Beliveau et al., 2017)
- 5-HT_4_: (Beliveau et al., 2017)
- 5-HT_6_: (Radhakrishnan et al., 2018)
- 5-HTT: (Beliveau et al., 2017)

Noradrenaline:

- NET: (Ding et al., 2010)

Acetylcholine:

- α4β2: (Hillmer et al., 2016)
- M_1_: (Naganawa et al., 2021)
- VAChT: (Aghourian et al., 2017; Bedard et al., 2019; Hansen et al., 2022)

GABA:

- GABA_A/BZ_: (Norgaard et al., 2021)

Glutamate:

- mGluR5: (DuBois et al., 2016; Hansen et al., 2022; Smart et al., 2019)

Histamine:

- H_3_: (Gallezot et al., 2017)

Endocannabinoids:

- CB_1_: (Normandin et al., 2015)

Opioids:

- μ: (Kantonen et al., 2020)

**Supplementary Table 1.** Demographics of the different cohorts.

| **Cohort** | **Group** | **Number of participants** | **Age (SD)** | **Sex, n (% men)** | **MoCA** | **MDS-UPDRS-III** |
| --- | --- | --- | --- | --- | --- | --- |
| Paris | iRBD | 59 | 67.4 ± 6.2 | 52 (88) | 26.8 ± 2.8 | 8.5 ± 6.7 |
| Paris | Controls | 57 | 65.9 ± 8.5 | 45 (79) | 27.5 ± 2.2 | 4.0 ± 4.8 |
| Montreal | iRBD | 48 | 65.8 ± 6.4 | 37 (77) | 25.9 ± 2.7 | 4.3 ± 3.6^a^ |
| Montreal | Controls | 35 | 65.0 ± 7.1 | 21 (60) | 28.2 ± 1.4 | - |
| Sydney | iRBD | 30 | 68.4 ± 7.2 | 27 (90) | 26.4 ± 3.3 | 11.6 ± 7.5 |
| Sydney | Controls | 26 | 70.8 ± 6.2 | 17 (65) | - | - |
| Aarhus | iRBD | 18 | 68.7 ± 8.2 | 15 (83) | 27.1 ± 2.0 | 1.1 ± 1.5 |
| Aarhus | Controls | 20 | 67.9 ± 5.8 | 15 (75) | 27.1 ± 2.1 | - |
| PPMI cohort | iRBD | 27 | 70.9 ± 5.3 | 21 (78) | 25.2 ± 4.5 | 5.1 ± 5.6 |
| PPMI cohort | Controls | 123 | 66.1 ± 8.2 | 97 (79) | 28.2 ± 1.1 | 1.4 ± 2.5 |

Data are presented as mean ± SD.
^a^Fahn & Elton UPDRS-III version (Fahn, Elton, & Members of the UPDRS Development Committee, 1987).

iRBD = isolated REM sleep behaviour disorder; MDS = Movement Disorders Society; MoCA = Montreal Cognitive Assessment; PPMI = Parkinson’s Progression Markers Initiative; SD = standard deviation; UPDRS-III = Unified Parkinson’s Disease Rating Scale, motor examination.

**Supplementary Table 2.** Associations between cell type gene expression and cortical thinning in iRBD.

| **Cell type** | **Cortical thickness** | | | |
| --- | --- | --- | --- | --- |
|  | *r* | *p*_original_ | *p*_spatial_ | *p*_random_ |
| Astrocyte | 0.08 | 0.64 | 0.39 | 0.32 |
| Endothelial cell | 0.01 | 0.95 | 0.48 | 0.47 |
| Excitatory neuron | -0.26 | 0.13 | 0.10 | 0.06 |
| Inhibitory neuron | 0.04 | 0.83 | 0.41 | 0.41 |
| Microglia | -0.03 | 0.87 | 0.44 | 0.44 |
| Oligodendrocyte | -0.08 | 0.64 | 0.33 | 0.33 |
| OPC | 0.18 | 0.30 | 0.28 | 0.14 |

Spearman’s correlation coefficients between regional cortical thickness *W*-scores and the average regional expression of genes associated with each cell type. No correlations survived the Bonferroni-corrected threshold (*p*<0.0071 for 7 comparisons).

iRBD = isolated REM sleep behavior disorder; OPC = oligodendrocyte precursor cells; REM = rapid eye movement.

**Supplementary Table 3.** Enriched biological processes using GOrilla.

| **GO Term** | **Description** | ***P*-value** | **FDR q-value** | **Enrich-ment** | ***N*** | ***B*** | ***n*** | ***b*** |
| --- | --- | --- | --- | --- | --- | --- | --- | --- |
| **Cortical thickness** | | | | | | | | |
| *Negatively weighted (more expressed) with greater cortical thinning* | | | | | | | | |
| GO:0016241 | regulation of macroautophagy | 9.53E-7 | 6.94E-3 | 4.32 | 13979 | 165 | 314 | 16 |
| GO:0009894 | regulation of catabolic process | 8.97E-7 | 1.31E-2 | 2.17 | 13979 | 902 | 314 | 44 |
| *Positively weighted (less expressed) with greater cortical thinning* | | | | | | | | |
| GO:0008380 | RNA splicing | 1.79E-12 | 8.71E-9 | 5.44 | 13979 | 346 | 193 | 26 |
| GO:0000375 | RNA splicing, via transesterification reactions | 7.12E-8 | 6.48E-5 | 4.91 | 13979 | 251 | 193 | 17 |
| GO:0006397 | mRNA processing | 2.77E-10 | 5.05E-7 | 4.52 | 13979 | 401 | 193 | 25 |
| GO:0006396 | RNA processing | 5.27E-12 | 1.54E-8 | 3.56 | 13979 | 773 | 193 | 38 |
| GO:0016071 | mRNA metabolic process | 1.15E-8 | 1.28E-5 | 3.42 | 13979 | 593 | 193 | 28 |
| GO:0016070 | RNA metabolic process | 4.86E-12 | 1.77E-8 | 2.77 | 13979 | 1358 | 193 | 52 |
| GO:0045944 | positive regulation of transcription by RNA polymerase II | 2.1E-7 | 1.39E-4 | 2.61 | 13979 | 943 | 193 | 34 |
| GO:0090304 | nucleic acid metabolic process | 3.02E-14 | 4.39E-10 | 2.59 | 13979 | 1875 | 193 | 67 |
| GO:0051254 | positive regulation of RNA metabolic process | 8.67E-9 | 1.05E-5 | 2.43 | 13979 | 1372 | 193 | 46 |
| GO:0045893 | positive regulation of transcription, DNA-templated | 9.96E-8 | 8.06E-5 | 2.36 | 13979 | 1288 | 193 | 42 |
| **Cortical surface area** | | | | | | | | |
| *Negatively weighted (less expressed) with greater cortical surface area* | | | | | | | | |
| GO:0071805 | potassium ion transmembrane transport | 1.24E-10 | 4.53E-7 | 5.04 | 13979 | 126 | 506 | 23 |
| GO:0071804 | cellular potassium ion transport | 1.24E-10 | 3.62E-7 | 5.04 | 13979 | 126 | 506 | 23 |
| GO:0006813 | potassium ion transport | 2.81E-10 | 6.81E-7 | 4.85 | 13979 | 131 | 506 | 23 |
| GO:0098662 | inorganic cation transmembrane transport | 2.33E-11 | 1.7E-7 | 3.05 | 13979 | 408 | 506 | 45 |
| GO:0030001 | metal ion transport | 2,00E-12 | 2.92E-8 | 2.99 | 13979 | 471 | 506 | 51 |
| GO:0098660 | inorganic ion transmembrane transport | 2.59E-11 | 1.26E-7 | 2.87 | 13979 | 472 | 506 | 49 |
| GO:0015672 | monovalent inorganic cation transport | 2.13E-7 | 2.59E-4 | 2.87 | 13979 | 289 | 506 | 30 |
| GO:0098655 | cation transmembrane transport | 6.89E-10 | 1.43E-6 | 2.75 | 13979 | 452 | 506 | 45 |
| GO:0006812 | cation transport | 1.98E-9 | 3.21E-6 | 2.45 | 13979 | 587 | 506 | 52 |
| GO:0034220 | ion transmembrane transport | 1.68E-9 | 3.05E-6 | 2.34 | 13979 | 673 | 506 | 57 |
| *Positively weighted (more expressed) with greater cortical surface area* | | | | | | | | |
| GO:0010273 | detoxification of copper ion | 7.52E-11 | 2.74E-7 | 24.92 | 13979 | 11 | 408 | 8 |
| GO:0061687 | detoxification of inorganic compound | 2.2E-10 | 4.58E-7 | 22.84 | 13979 | 12 | 408 | 8 |
| GO:0071294 | cellular response to zinc ion | 1.67E-8 | 1.52E-5 | 15.23 | 13979 | 18 | 408 | 8 |
| GO:0006882 | cellular zinc ion homeostasis | 1.25E-8 | 1.21E-5 | 12.85 | 13979 | 24 | 408 | 9 |
| GO:0055069 | zinc ion homeostasis | 2.18E-9 | 2.64E-6 | 12.69 | 13979 | 27 | 408 | 10 |
| GO:0071280 | cellular response to copper ion | 1.1E-7 | 6.4E-5 | 12.46 | 13979 | 22 | 408 | 8 |
| GO:0046688 | response to copper ion | 4.9E-9 | 5.49E-6 | 11.81 | 13979 | 29 | 408 | 10 |
| GO:0071276 | cellular response to cadmium ion | 1.24E-9 | 1.81E-6 | 11.42 | 13979 | 33 | 408 | 11 |
| GO:0007159 | leukocyte cell-cell adhesion | 1.2E-7 | 6.46E-5 | 8.79 | 13979 | 39 | 408 | 10 |
| GO:0042116 | macrophage activation | 6.58E-7 | 2.23E-4 | 8.57 | 13979 | 36 | 408 | 9 |

The top 10 biological process terms enriched in the genes predicting cortical thickness and surface *W*-scores in iRBD ranked based on the enrichment ratio. *N* represents the total number of genes, *B* the total number of genes associated with a specific GO term, *n* the number of genes in the target list, and *b* the number of genes in the intersection. The enrichment score was calculated as (*b*/*n*)/(*B*/*N*).

FDR = false discovery rate; GO = Gene Ontology; iRBD = isolated REM sleep behavior disorder; REM = rapid eye movement.

**Supplementary Table 4.** Enriched cellular components using GOrilla.

| **GO Term** | **Description** | ***P*-value** | **FDR q-value** | **Enrich-ment** | ***N*** | ***B*** | ***n*** | ***b*** |
| --- | --- | --- | --- | --- | --- | --- | --- | --- |
| **Cortical thickness** | | | | | | | | |
| *Negatively weighted (more expressed) with greater cortical thinning* | | | | | | | | |
| GO:0044455 | mitochondrial membrane part | 2.81E-7 | 8.84E-5 | 4.22 | 13979 | 190 | 314 | 18 |
| GO:0098798 | mitochondrial protein complex | 1.61E-7 | 6.08E-5 | 3.97 | 13979 | 224 | 314 | 20 |
| GO:0044429 | mitochondrial part | 7.59E-7 | 1.79E-4 | 2.16 | 13979 | 927 | 314 | 45 |
| GO:1902494 | catalytic complex | 1.45E-8 | 9.11E-6 | 2.14 | 13979 | 1227 | 314 | 59 |
| GO:0005739 | mitochondrion | 4.72E-7 | 1.27E-4 | 2.05 | 13979 | 1129 | 314 | 52 |
| GO:0032991 | protein-containing complex | 1.01E-8 | 9.49E-6 | 1.48 | 13979 | 4465 | 314 | 148 |
| GO:0005829 | cytosol | 3.06E-8 | 1.44E-5 | 1.46 | 13979 | 4452 | 314 | 146 |
| GO:0044444 | cytoplasmic part | 7.22E-9 | 1.36E-5 | 1.27 | 13979 | 8087 | 314 | 230 |
| *Positively weighted (less expressed) with greater cortical thinning* | | | | | | | | |
| GO:0044451 | nucleoplasm part | 3.64E-8 | 1.72E-5 | 2.65 | 13979 | 1010 | 193 | 37 |
| GO:0005654 | nucleoplasm | 1.02E-8 | 6.38E-6 | 1.78 | 13979 | 3298 | 193 | 81 |
| GO:0044428 | nuclear part | 7.05E-10 | 6.64E-7 | 1.71 | 13979 | 4106 | 193 | 97 |
| GO:0005634 | nucleus | 5.29E-10 | 9.98E-7 | 1.68 | 13979 | 4343 | 193 | 101 |
| **Cortical surface area** | | | | | | | | |
| *Negatively weighted (less expressed) with greater cortical surface area* | | | | | | | | |
| GO:0034703 | cation channel complex | 5.95E-9 | 5.61E-6 | 3.88 | 13979 | 178 | 506 | 25 |
| GO:1902495 | transmembrane transporter complex | 2.44E-9 | 4.61E-6 | 3.40 | 13979 | 252 | 506 | 31 |
| GO:0034702 | ion channel complex | 3.25E-8 | 1.53E-5 | 3.28 | 13979 | 236 | 506 | 28 |
| GO:1990351 | transporter complex | 9.89E-9 | 6.22E-6 | 3.21 | 13979 | 267 | 506 | 31 |
| *Positively weighted (more expressed) with greater cortical surface area* | | | | | | | | |
| GO:0009986 | cell surface | 9.83E-7 | 6.18E-4 | 2.62 | 13979 | 405 | 408 | 31 |
| GO:0044459 | plasma membrane part | 4.1E-8 | 7.74E-5 | 1.69 | 13979 | 2027 | 408 | 100 |
| GO:0005886 | plasma membrane | 5.99E-8 | 5.64E-5 | 1.50 | 13979 | 3165 | 408 | 139 |

The top 10 cellular component terms enriched in the genes predicting cortical thickness and surface area *W*-scores in iRBD ranked based on the enrichment ratio. *N* represents the total number of genes, *B* the total number of genes associated with a specific GO term, *n* the number of genes in the target list, and *b* the number of genes in the intersection. The enrichment score was calculated as (*b*/*n*)/(*B*/*N*).

FDR = false discovery rate; GO = Gene Ontology; iRBD = isolated REM sleep behavior disorder; REM = rapid eye movement.

**Supplementary Table 5.** GSEA of the genes predicting cortical surface area changes in iRBD.

| **GO identifier** | **GO term** | **Gene set size** | **Number of leading edge IDs** | **Enrichment score** | **Normalized enrichment score** | **FDR *P*-value** |
| --- | --- | --- | --- | --- | --- | --- |
| ***Negatively weighted (less expressed) genes*** | | | | | | |
| *Biological processes* | | | | | | |
| GO:0006813 | potassium ion transport | 188 | 66 | -0.379 | -1.810 | 0.269 |
| GO:0006984 | ER-nucleus signaling pathway | 40 | 17 | -0.470 | -1.729 | 0.307 |
| GO:0000959 | mitochondrial RNA metabolic process | 40 | 16 | -0.474 | -1.705 | 0.260 |
| GO:0034067 | protein localization to Golgi apparatus | 24 | 14 | -0.532 | -1.701 | 0.205 |
| GO:0044872 | lipoprotein localization | 11 | 3 | -0.635 | -1.626 | 0.331 |
| GO:0034765 | regulation of ion transmembrane transport | 353 | 108 | -0.313 | -1.598 | 0.349 |
| GO:0008214 | protein dealkylation | 31 | 12 | -0.475 | -1.593 | 0.314 |
| GO:0140115 | export across plasma membrane | 24 | 9 | -0.490 | -1.586 | 0.294 |
| GO:0097503 | sialylation | 18 | 10 | -0.511 | -1.523 | 0.400 |
| GO:0015931 | nucleobase-containing compound transport | 217 | 64 | -0.309 | -1.485 | 0.414 |
| *Cellular components* | | | | | | |
| GO:0016234 | inclusion body | 71 | 29 | -0.425 | -1.750 | 0.130 |
| GO:1990351 | transporter complex | 262 | 62 | -0.335 | -1.666 | 0.160 |
| GO:0009295 | nucleoid | 42 | 16 | -0.446 | -1.627 | 0.156 |
| GO:0031519 | polycomb group protein complex | 42 | 13 | -0.435 | -1.599 | 0.151 |
| GO:0070971 | endoplasmic reticulum exit site | 20 | 8 | -0.496 | -1.520 | 0.228 |
| GO:0044298 | cell body membrane | 23 | 9 | -0.474 | -1.519 | 0.192 |
| GO:0098982 | GABAergic synapse | 69 | 19 | -0.367 | -1.458 | 0.259 |
| GO:0048475 | coated membrane | 90 | 38 | -0.327 | -1.384 | 0.398 |
| GO:1903293 | phosphatase complex | 47 | 15 | -0.359 | -1.349 | 0.456 |
| GO:0098687 | chromosomal region | 273 | 111 | -0.260 | -1.274 | 0.536 |
| *Disease-related terms* | | | | | | |
| *DisGeNET* |  |  |  |  |  |  |
| C0004134 | Ataxia | 13 | 7 | -0.82204 | -2.2706 | **0.0016** |
| ***Positively weighted (more expressed) genes*** | | | | | | |
| *Biological processes* | | | | | | |
| GO:0002269 | leukocyte activation involved in inflammatory response | 22 | 11 | 0.635 | 2.044 | **0.023** |
| GO:0006968 | cellular defense response | 32 | 13 | 0.570 | 2.010 | **0.024** |
| GO:0006959 | humoral immune response | 116 | 38 | 0.437 | 1.977 | **0.025** |
| GO:0055076 | transition metal ion homeostasis | 113 | 28 | 0.429 | 1.957 | **0.023** |
| GO:0010573 | vascular endothelial growth factor production | 23 | 11 | 0.597 | 1.940 | **0.023** |
| GO:0070661 | leukocyte proliferation | 183 | 60 | 0.401 | 1.938 | **0.020** |
| GO:0070972 | protein localization to endoplasmic reticulum | 135 | 56 | 0.416 | 1.938 | **0.018** |
| GO:0150076 | neuroinflammatory response | 32 | 13 | 0.550 | 1.920 | **0.019** |
| GO:0036230 | granulocyte activation | 386 | 111 | 0.347 | 1.837 | **0.029** |
| *Cellular components* | | | | | | |
| GO:0042611 | MHC protein complex | 18 | 11 | 0.717 | 2.130 | **0.002** |
| GO:0097386 | glial cell projection | 21 | 8 | 0.635 | 2.038 | **0.003** |
| GO:1905360 | GTPase complex | 24 | 12 | 0.626 | 2.029 | **0.002** |
| GO:0005766 | primary lysosome | 127 | 43 | 0.415 | 1.903 | **0.010** |
| GO:0030055 | cell-substrate junction | 377 | 108 | 0.346 | 1.807 | **0.021** |
| GO:0030667 | secretory granule membrane | 216 | 75 | 0.351 | 1.743 | **0.026** |
| GO:0044445 | cytosolic part | 226 | 80 | 0.347 | 1.733 | **0.025** |
| GO:0005775 | vacuolar lumen | 136 | 39 | 0.371 | 1.715 | **0.026** |
| GO:0098862 | cluster of actin-based cell projections | 101 | 27 | 0.376 | 1.693 | **0.025** |
| GO:0101002 | ficolin-1-rich granule | 155 | 39 | 0.348 | 1.657 | **0.027** |
| *Disease-related terms* | | | | | | |
| *PharmGKB* |  |  |  |  |  |  |
| PA445299 | Aggressive periodontitis | 24 | 13 | 0.66089 | 2.1530 | **0.0289** |
| PA444530 | Hyperlipoproteinemias | 20 | 8 | 0.68962 | 2.1460 | **0.0151** |

The top 10 biological process and cellular component terms enriched in the genes predicting cortical surface area in iRBD are reported ranked based on the normalized enrichment score. Bold values represent the terms that were significant enriched in the gene set predicting surface area after applying FDR correction. The significant disease-related gene terms predicting surface area are also listed.

ER = endoplasmic reticulum; FDR = false discovery rate; GO = Gene Ontology; GSEA = gene set enrichment analysis; iRBD = isolated REM sleep behavior disorder; MHC = major histocompatibility complex; REM = rapid eye movement.

**Supplementary Table 6.** Associations between cell type gene expression and cortical surface area in iRBD.

| **Cell type** | **Cortical surface area** | | | |
| --- | --- | --- | --- | --- |
|  | *r* | *p*_original_ | *p*_spatial_ | *p*_random_ |
| Astrocyte | 0.61 | **0.0002** | **0.0011** | **<0.0001** |
| Endothelial cell | -0.08 | 0.63 | 0.34 | 0.32 |
| Excitatory neuron | -0.37 | 0.031 | 0.028 | 0.016 |
| Inhibitory neuron | -0.31 | 0.07 | 0.040 | 0.034 |
| Microglia | 0.52 | **0.0019** | **0.0053** | **0.0010** |
| Oligodendrocyte | -0.06 | 0.74 | 0.37 | 0.37 |
| OPC | 0.53 | **0.0015** | **0.0049** | **0.0007** |

Spearman’s correlation coefficients between regional cortical surface area *W*-scores and the average regional expression of genes associated with each cell type. Bold values represent the original correlations that survived the Bonferroni-corrected threshold (*p*<0.0071 for 7 comparisons) and that were still significant when tested against spatially constrained null models.

iRBD = isolated REM sleep behavior disorder; OPC = oligodendrocyte precursor cells; REM = rapid eye movement.

**Supplementary Figure 1.** Patterns of gene expression underlying cortical surface area changes in iRBD.

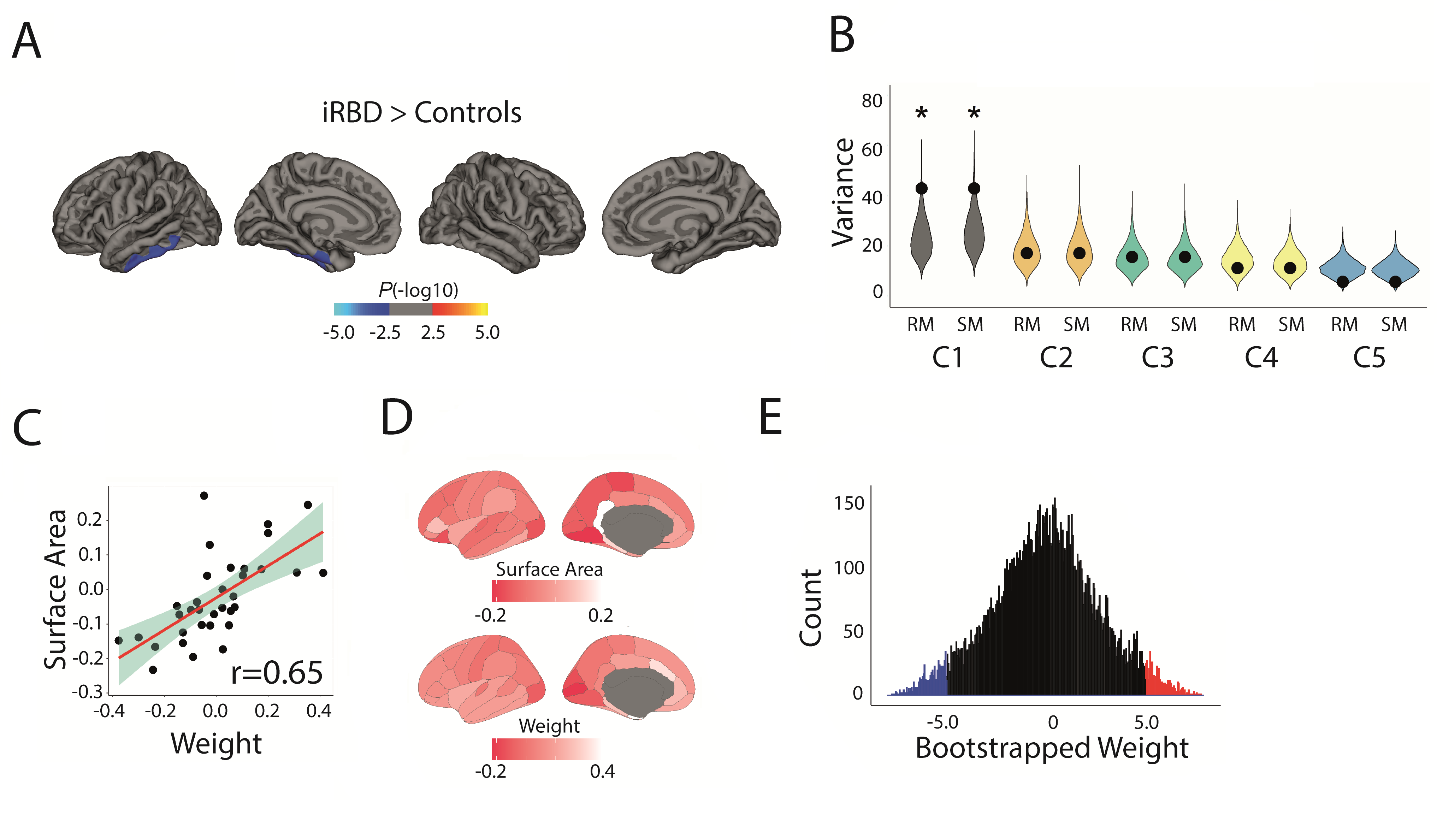

**(A)** Vertex-wise patterns showing the significant changes in cortical surface area in iRBD patients compared to controls. **(B)** Violin plots showing the percentage of variance in cortical surface area *W*-scores explained by gene expression; the dot represents the empirical variance, and the asterisk indicates the components that were significant against random and spatial null models. **(C)** Scatterplots of the associations between surface area *W*-scores and the regional weights of the first components. **(D)** Brain renderings of the surface area *W*-scores and the regional weights of the first components. **(E)** Density plots of each gene’s bootstrapped weight on the first components; gene set enrichment analysis was performed on all genes, whereas over-representation analysis was performed on genes with bootstrap ratios ± 5.0.

C = component; iRBD = isolated REM sleep behavior disorder; REM = rapid eye movement; RM = random null models; SM = spatial null models.

**Supplementary Figure 2.** Enrichment analyses of the genes associated with cortical surface area changes in iRBD.

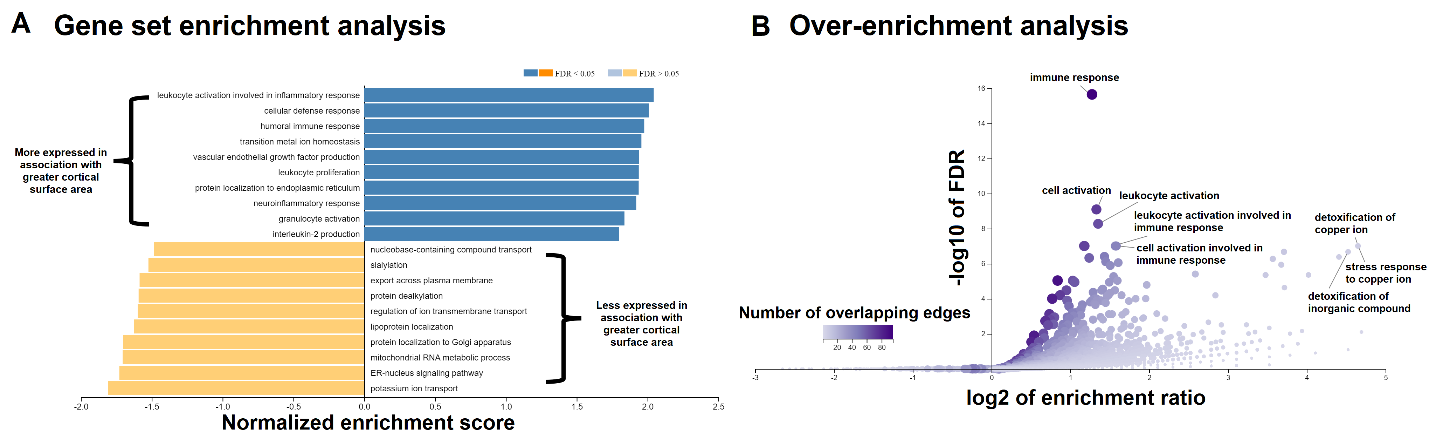

**(A)** The top 10 biological process terms from the GO Consortium knowledge base that are enriched in the positively and negatively weighted gene sets predicting cortical surface area changes in iRBD. Terms are ranked based on the normalized enrichment score; darker colored bars present significantly enriched terms after FDR correction. **(B)** Volcano plots of the over-representation analyses showing the biological process terms enriched in the genes most strongly associated with greater cortical surface area in iRBD (bootstrap ratio >5.0 for surface area). The color bar represents the number of overlapping edges for each gene category and the size of the dot represents the size of the gene category.

FDR = false discovery rate; GO = Gene Ontology; iRBD = isolated REM sleep behavior disorder; REM = rapid eye movement.

**Supplementary Figure 3.** Relationship between cell type gene expression and cortical surface area changes in iRBD.

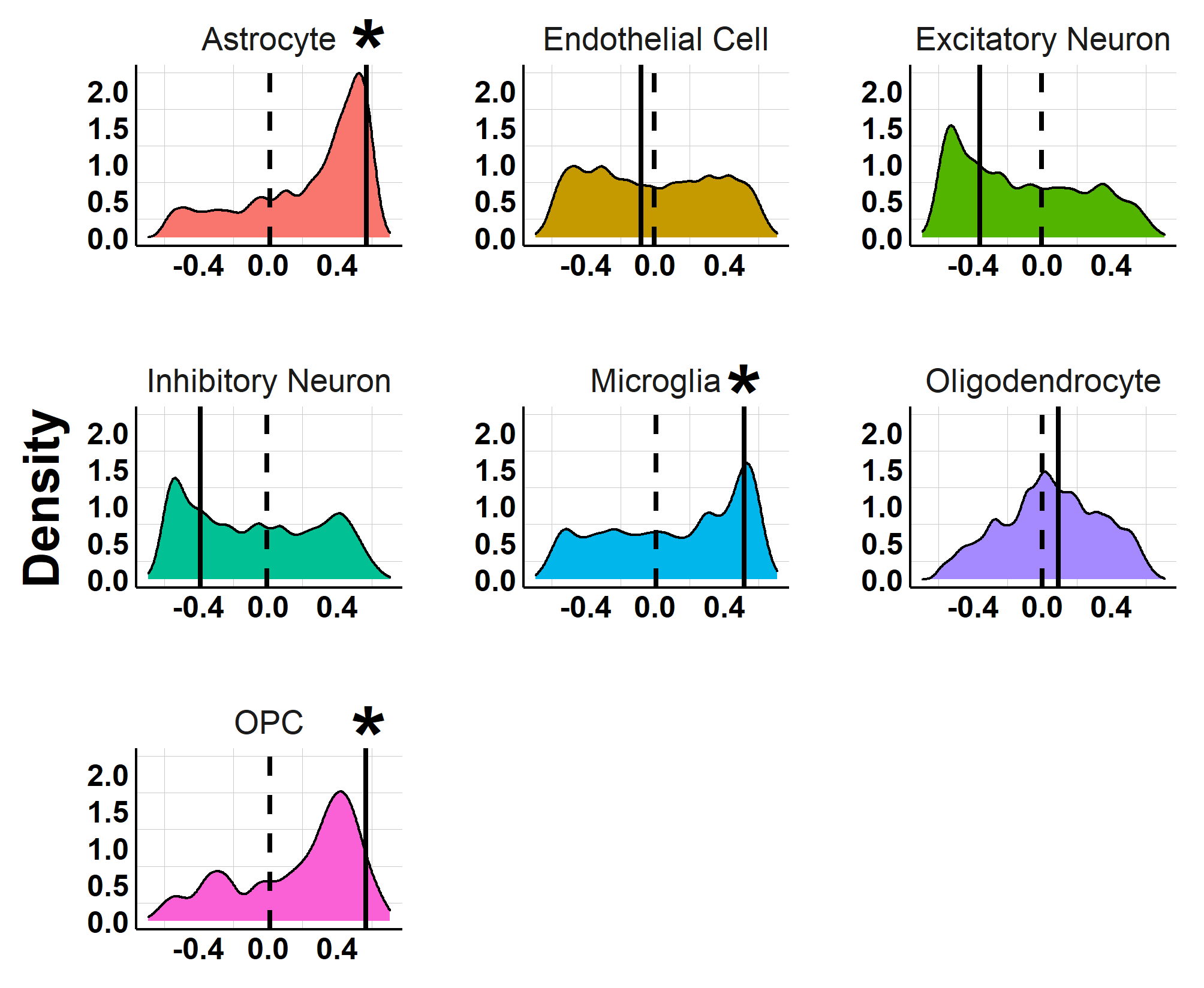

Density plots showing the distribution of the correlation coefficients between regional cortical surface area *W­*-scores and the average regional expression of genes associated with each of the 7 cell types. The straight line represents the empirical correlation, and the dashed line represents the average correlation coefficient observed in sets of 10,000 spatially constrained null models. The asterisk indicates the associations significant under the Bonferroni-corrected threshold of *p*<0.007 that remained significant when tested against null models.

iRBD = isolated REM sleep behavior disorder; OPC = oligodendrocyte precursor cells; REM = rapid eye movement.

**Supplementary Figure 4.** The connectome constrains cortical surface changes in iRBD.

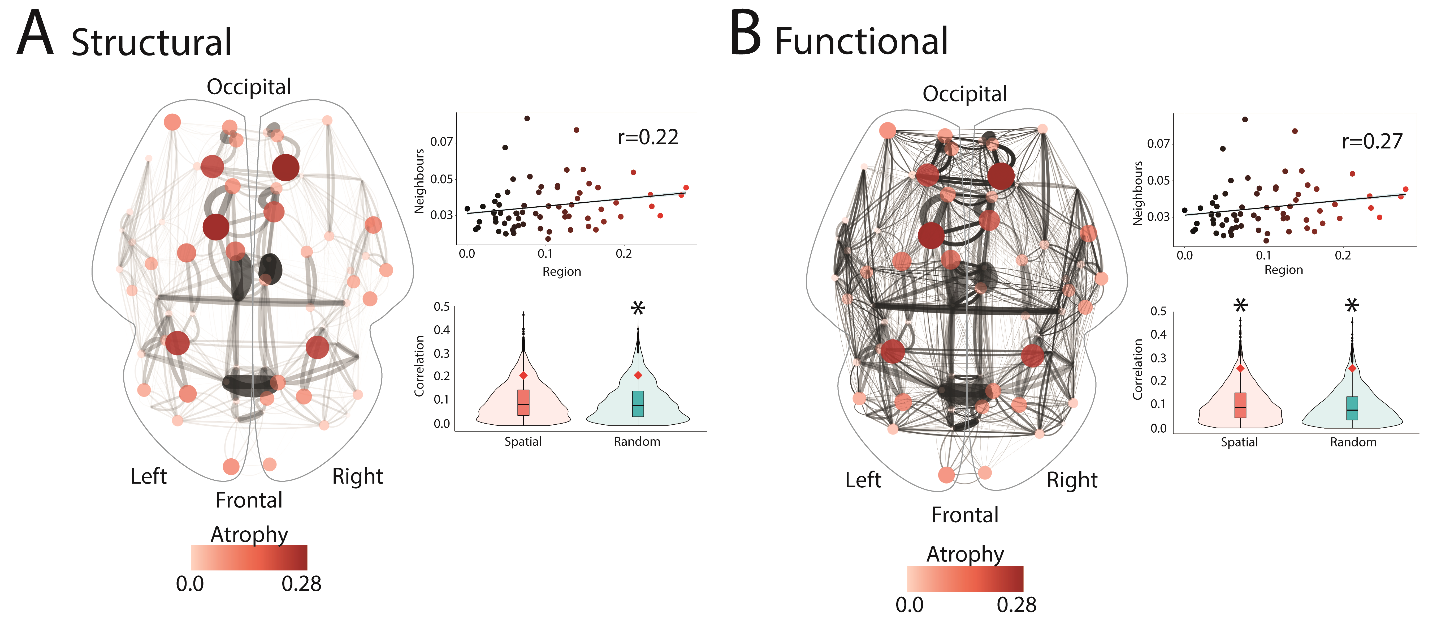

Brain renderings showing the associations between the deviation in cortical surface area *W*-scores in iRBD and **(A)** structural and **(B)** functional connectivity. The edge thickness represents the interregional connection strength, whereas the node size and color represent the local deviation in the *W*-score in iRBD compared to controls (i.e., the larger and redder, the more local surface area deviated from controls). The scatterplots show the associations between the deviations in *W*-scores and the average *W*-scores observed in structural or functional neighbors. The violin plots show the empirical correlation against sets of 10,000 correlations generated from spatial and random null models. The asterisk indicates associations that were significant against null models.

iRBD = isolated REM sleep behavior disorder; REM = rapid eye movement.

**Supplementary Figure 5.** Cortical surface area changes in iRBD map onto regional tracer density.

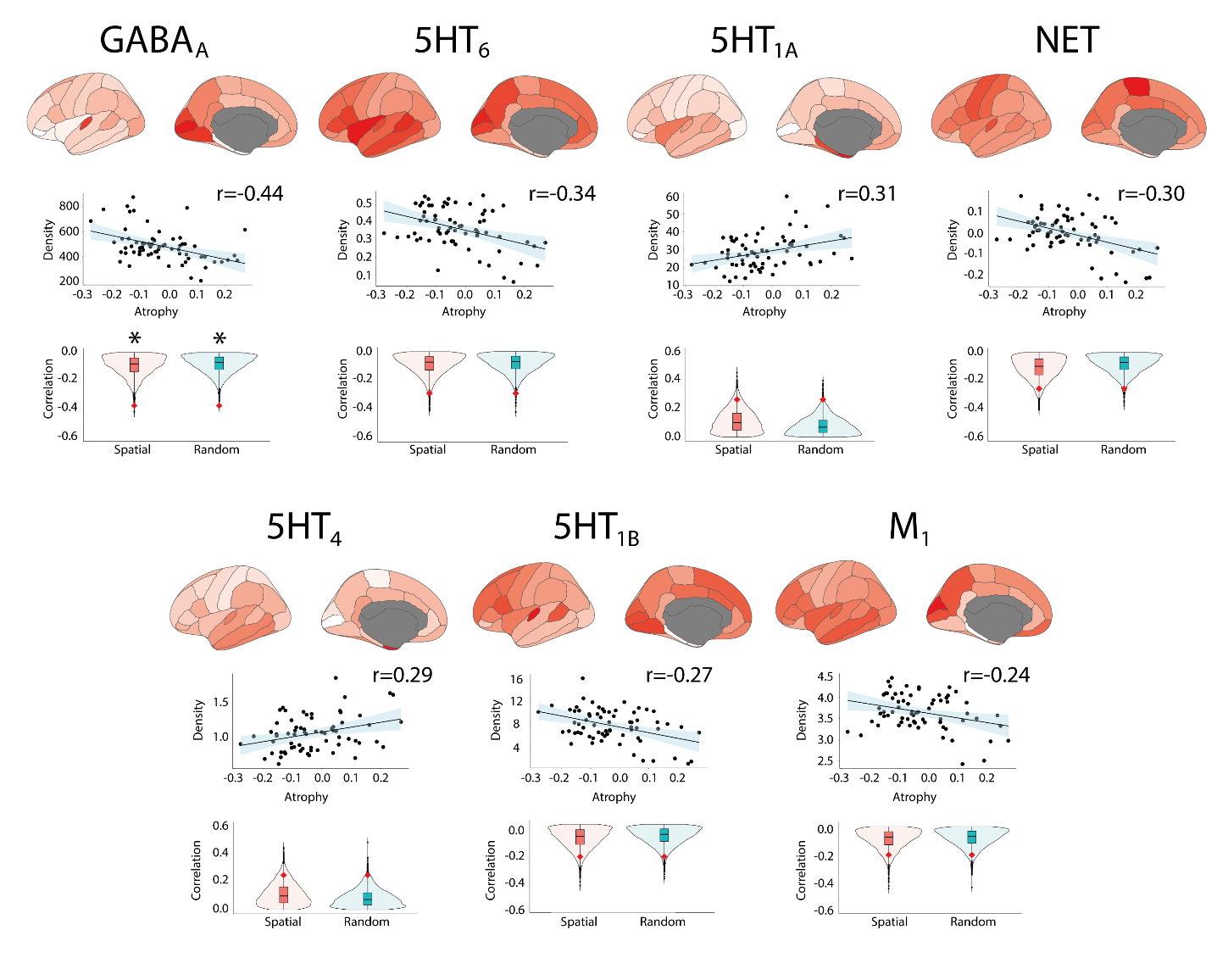

Brain renderings and scatterplots showing the tracer density maps of the receptors, transporters, and binding sites associated with cortical surface area *W*-scores in iRBD. The violin plots show the empirical correlations tested against distributions of correlations from sets of spatial and random null models. The asterisk indicates the significant associations after Bonferroni correction.

iRBD = isolated REM sleep behavior disorder; REM = rapid eye movement.

**Supplementary Figure 6.** Cortical surface area changes in iRBD map onto specific brain systems.

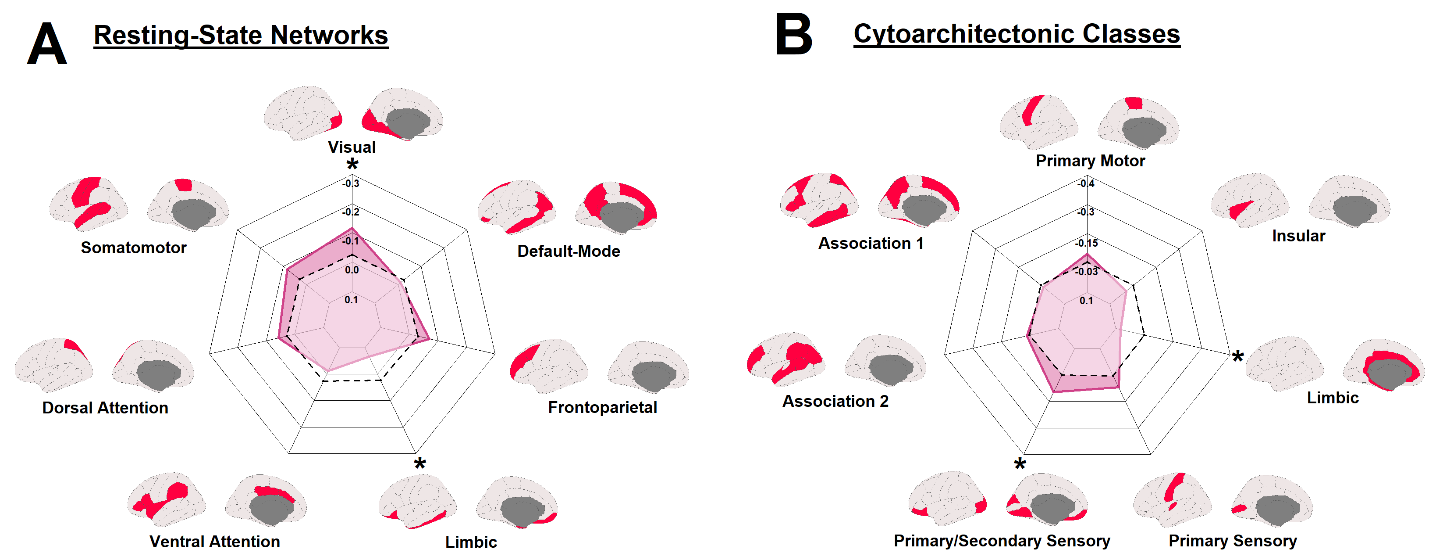

Radar charts showing the correlation between cortical surface area *W*-scores in iRBD and **(A)** resting-state networks and **(B)** cytoarchitectonic classes. The regular line represents the empirical correlations, and the dashed line represents the average correlation observed in sets of 10,000 spatial null models. The asterisk indicates the networks and classes where the observed spatial correlation was significantly different from the null correlation.

iRBD = isolated REM sleep behavior disorder; REM = rapid eye movement.

**Supplementary Figure 7.** Cortical surface area changes in iRBD map relate to distinct cognitive processes.

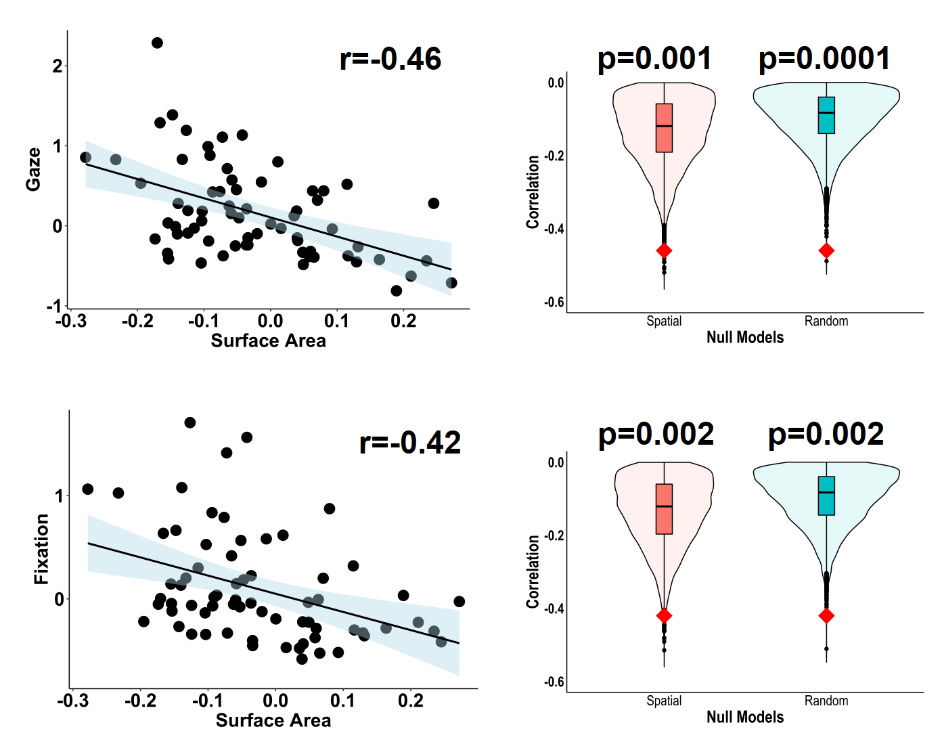

Scatterplots showing the associations between cortical surface area *W*-scores in iRBD and the functional correlates associated with cognitive processes. The violin plots show the empirical correlations compared against the distributions of correlations from sets of 10,000 spatial and random null models. Of the 123 cognitive processes tested, only 2 were significant after correcting for Bonferroni.

iRBD = isolated REM sleep behavior disorder; REM = rapid eye movement.

**REFERENCES**

Aghourian, M., Legault-Denis, C., Soucy, J. P., Rosa-Neto, P., Gauthier, S., Kostikov, A., . . . Bedard, M. A. (2017). Quantification of brain cholinergic denervation in Alzheimer's disease using PET imaging with [(18)F]-FEOBV. *Mol Psychiatry, 22*(11), 1531-1538. doi:10.1038/mp.2017.183

Bedard, M. A., Aghourian, M., Legault-Denis, C., Postuma, R. B., Soucy, J. P., Gagnon, J. F., . . . Montplaisir, J. (2019). Brain cholinergic alterations in idiopathic REM sleep behaviour disorder: a PET imaging study with (18)F-FEOBV. *Sleep Med, 58*, 35-41. doi:10.1016/j.sleep.2018.12.020

Beliveau, V., Ganz, M., Feng, L., Ozenne, B., Hojgaard, L., Fisher, P. M., . . . Knudsen, G. M. (2017). A High-Resolution In Vivo Atlas of the Human Brain's Serotonin System. *J Neurosci, 37*(1), 120-128. doi:10.1523/JNEUROSCI.2830-16.2016

Ding, Y. S., Singhal, T., Planeta-Wilson, B., Gallezot, J. D., Nabulsi, N., Labaree, D., . . . Malison, R. T. (2010). PET imaging of the effects of age and cocaine on the norepinephrine transporter in the human brain using (S,S)-[(11)C]O-methylreboxetine and HRRT. *Synapse, 64*(1), 30-38. doi:10.1002/syn.20696

DuBois, J. M., Rousset, O. G., Rowley, J., Porras-Betancourt, M., Reader, A. J., Labbe, A., . . . Kobayashi, E. (2016). Characterization of age/sex and the regional distribution of mGluR5 availability in the healthy human brain measured by high-resolution [(11)C]ABP688 PET. *Eur J Nucl Med Mol Imaging, 43*(1), 152-162. doi:10.1007/s00259-015-3167-6

Dukart, J., Holiga, S., Chatham, C., Hawkins, P., Forsyth, A., McMillan, R., . . . Sambataro, F. (2018). Cerebral blood flow predicts differential neurotransmitter activity. *Sci Rep, 8*(1), 4074. doi:10.1038/s41598-018-22444-0

Fahn, S., Elton, R., & Members of the UPDRS Development Committee. (1987). Unified Parkinson's Disease Rating Scale. In S. Fahn, C. D. Marsden, D. B. Calne, & M. Goldstein (Eds.), *Recent developments in Parkinson's disease* (Vol. 2, pp. 153-163, 293-304). Florham Park, NJ: Macmillan Health Care Information.

Gallezot, J. D., Nabulsi, N., Neumeister, A., Planeta-Wilson, B., Williams, W. A., Singhal, T., . . . Carson, R. E. (2010). Kinetic modeling of the serotonin 5-HT(1B) receptor radioligand [(11)C]P943 in humans. *J Cereb Blood Flow Metab, 30*(1), 196-210. doi:10.1038/jcbfm.2009.195

Gallezot, J. D., Planeta, B., Nabulsi, N., Palumbo, D., Li, X., Liu, J., . . . Carson, R. E. (2017). Determination of receptor occupancy in the presence of mass dose: [(11)C]GSK189254 PET imaging of histamine H3 receptor occupancy by PF-03654746. *J Cereb Blood Flow Metab, 37*(3), 1095-1107. doi:10.1177/0271678X16650697

Hansen, J. Y., Shafiei, G., Markello, R. D., Smart, K., Cox, S. M. L., Nørgaard, M., . . . Misic, B. (2022). Mapping neurotransmitter systems to the structural and functional organization of the human neocortex. *bioRxiv*, 2021.2010.2028.466336. doi:10.1101/2021.10.28.466336

Hillmer, A. T., Esterlis, I., Gallezot, J. D., Bois, F., Zheng, M. Q., Nabulsi, N., . . . Cosgrove, K. P. (2016). Imaging of cerebral alpha4beta2* nicotinic acetylcholine receptors with (-)-[(18)F]Flubatine PET: Implementation of bolus plus constant infusion and sensitivity to acetylcholine in human brain. *Neuroimage, 141*, 71-80. doi:10.1016/j.neuroimage.2016.07.026

Kaller, S., Rullmann, M., Patt, M., Becker, G. A., Luthardt, J., Girbardt, J., . . . Sabri, O. (2017). Test-retest measurements of dopamine D1-type receptors using simultaneous PET/MRI imaging. *Eur J Nucl Med Mol Imaging, 44*(6), 1025-1032. doi:10.1007/s00259-017-3645-0

Kantonen, T., Karjalainen, T., Isojarvi, J., Nuutila, P., Tuisku, J., Rinne, J., . . . Nummenmaa, L. (2020). Interindividual variability and lateralization of mu-opioid receptors in the human brain. *Neuroimage, 217*, 116922. doi:10.1016/j.neuroimage.2020.116922

Marek, K., Chowdhury, S., Siderowf, A., Lasch, S., Coffey, C. S., Caspell-Garcia, C., . . . Parkinson's Progression Markers, I. (2018). The Parkinson's progression markers initiative (PPMI) - establishing a PD biomarker cohort. *Ann Clin Transl Neurol, 5*(12), 1460-1477. doi:10.1002/acn3.644

Naganawa, M., Nabulsi, N., Henry, S., Matuskey, D., Lin, S. F., Slieker, L., . . . Huang, Y. (2021). First-in-Human Assessment of (11)C-LSN3172176, an M1 Muscarinic Acetylcholine Receptor PET Radiotracer. *J Nucl Med, 62*(4), 553-560. doi:10.2967/jnumed.120.246967

Norgaard, M., Beliveau, V., Ganz, M., Svarer, C., Pinborg, L. H., Keller, S. H., . . . Knudsen, G. M. (2021). A high-resolution in vivo atlas of the human brain's benzodiazepine binding site of GABAA receptors. *Neuroimage, 232*, 117878. doi:10.1016/j.neuroimage.2021.117878

Normandin, M. D., Zheng, M. Q., Lin, K. S., Mason, N. S., Lin, S. F., Ropchan, J., . . . Huang, Y. (2015). Imaging the cannabinoid CB1 receptor in humans with [11C]OMAR: assessment of kinetic analysis methods, test-retest reproducibility, and gender differences. *J Cereb Blood Flow Metab, 35*(8), 1313-1322. doi:10.1038/jcbfm.2015.46

Radhakrishnan, R., Nabulsi, N., Gaiser, E., Gallezot, J. D., Henry, S., Planeta, B., . . . Matuskey, D. (2018). Age-Related Change in 5-HT6 Receptor Availability in Healthy Male Volunteers Measured with (11)C-GSK215083 PET. *J Nucl Med, 59*(9), 1445-1450. doi:10.2967/jnumed.117.206516

Sandiego, C. M., Gallezot, J. D., Lim, K., Ropchan, J., Lin, S. F., Gao, H., . . . Cosgrove, K. P. (2015). Reference region modeling approaches for amphetamine challenge studies with [11C]FLB 457 and PET. *J Cereb Blood Flow Metab, 35*(4), 623-629. doi:10.1038/jcbfm.2014.237

Savli, M., Bauer, A., Mitterhauser, M., Ding, Y. S., Hahn, A., Kroll, T., . . . Lanzenberger, R. (2012). Normative database of the serotonergic system in healthy subjects using multi-tracer PET. *Neuroimage, 63*(1), 447-459. doi:10.1016/j.neuroimage.2012.07.001

Smart, K., Cox, S. M. L., Scala, S. G., Tippler, M., Jaworska, N., Boivin, M., . . . Leyton, M. (2019). Sex differences in [(11)C]ABP688 binding: a positron emission tomography study of mGlu5 receptors. *Eur J Nucl Med Mol Imaging, 46*(5), 1179-1183. doi:10.1007/s00259-018-4252-4

Smith, C. T., Crawford, J. L., Dang, L. C., Seaman, K. L., San Juan, M. D., Vijay, A., . . . Samanez-Larkin, G. R. (2019). Partial-volume correction increases estimated dopamine D2-like receptor binding potential and reduces adult age differences. *J Cereb Blood Flow Metab, 39*(5), 822-833. doi:10.1177/0271678X17737693
